# Recovery of serum testosterone levels is an accurate predictor of survival from COVID-19 in male patients

**DOI:** 10.1101/2021.06.29.21259693

**Authors:** Emily Toscano-Guerra, Mónica Martínez Gallo, Iria Arrese-Muñoz, Anna Giné, Noelia Díaz, Pablo Gabriel-Medina, Mar Riveiro-Barciela, Moisés Labrador-Horrillo, Fernando Martinez-Valle, Manuel Hernández-González, Ricardo Pujol Borrell, Francisco Rodríguez-Frias, Roser Ferrer, Timothy M. Thomson, Rosanna Paciucci

**Affiliations:** Biochemistry Service, Vall d’Hebron Hospital, Barcelona, Spain; Cell Signaling and Cancer Progression Laboratory, Vall d’Hebron Institute of Research, Barcelona, Spain; Immunology Division, Vall d’Hebron Hospital, Barcelona, Spain; Diagnostic Immunology Research Group, Vall d’Hebron Research Institute (VHIR), Barcelona, Spain; Department of Cell Biology, Physiology and Immunology, Autonomous University of Barcelona (UAB), Barcelona, Spain; Internal Medicine Service, Vall d’Hebron Hospital, Barcelona, Spain; Barcelona Institute for Molecular Biology, National Science Council (IBMB-CSIC), Barcelona, Spain; Networked Center for Hepatic and Digestive Diseases (CIBER-EHD), Instituto Nacional de la Salud Carlos III, Madrid, Spain

**Keywords:** COVID-19, survival, longitudinal, testosterone, immune phenotype

## Abstract

Infection with SARS-CoV-2 portends a broad range of outcomes, from a majority of asymptomatic cases or mild clinical courses to a lethal disease. Robust correlates of severe COVID-19 include old age, male sex, poverty and co-morbidities such as obesity, diabetes or cardiovascular disease. A precise knowledge is still lacking of the molecular and biological mechanisms that may explain the association of severe disease with male sex. Here, we show that testosterone trajectories are highly accurate individual predictors (AUC of ROC = 0.928, *p* < 0.0001) of survival in male COVID-19 patients. Longitudinal determinations of blood levels of luteinizing hormone (LH) and androstenedione suggest an early modest inhibition of the central LH-androgen biosynthesis axis in a majority of patients, followed by either full recovery in survivors or a peripheral failure in lethal cases. Moreover, failure to reinstate physiological testosterone levels was associated with evidence of impaired T helper differentiation and decrease of non-classical monocytes. The strong association of recovery or failure to reinstate testosterone levels with survival or death from COVID-19 in male patients is suggestive of a significant role of testosterone status in the immune responses to COVID-19.

## Introduction

The COVID-19 pandemic caused by the new SARS-CoV-2 virus is characterized by a diversity of clinical manifestations, including exacerbated inflammatory states accompanied with tissue and organ destruction beyond direct cytopathic effects of SARS-CoV-2. From the outset of the pandemic, it became clear that, while men and women present a similar prevalence of infection^1,2^, a higher risk of severe disease and death is significantly associated with male sex^2,3^. Similar observations come from studies of outbreaks of pathogenic coronaviruses with SARS-CoV and the Middle East respiratory syndrome (MERS-CoV), or other viral respiratory infections^4,5^. Multiple factors have been considered to explain the sex disparity observed in the development of severe COVID-19, including differential biological and pathophysiological impacts of age and comorbidities such as cardiovascular disease, high blood pressure, chronic obstructive pulmonary disease (COPD), diabetes, obesity or active cancer^6^. Sexual differences in disease severity are also observed in racial and ethnic minority groups, disproportionately affected by COVID-19^6^.

The underlying mechanisms that may account for these differences are not completely understood. As the main cellular receptor for SARS-CoV-2, ACE2, and the major viral fusogenic membrane-associated protease, TMPRSS2, are under transcriptional regulation by androgens^7^, it had been predicted that men would present a higher propensity of infection by SARS-CoV-2 and to develop more severe disease than women^7,8^. However, men and women show comparable risks of infection^1,2^ and observational studies of male COVID-19 patients under androgen-deprivation therapy have yielded contrasting results with regards to risk of developing severe COVID-19 (ref. 9,10). Contrariwise, there is growing evidence that severe COVID-19 in male patients is accompanied with diminished levels of circulating testosterone^11-13^, suggesting a critical role for androgens^11,14^ in preventing the innate and/or adaptive immune dysfunctions that lead to the development of severe forms of the disease^15-17^.

The sexual dimorphism of immune responses to pathogens has been long recognised^18^, pointing to women having stronger antiviral mechanisms, stronger T-regulatory cells, higher numbers of group 2 innate lymphoid cells (ILC2), and superior immune-mediated tissue repair capacities as compared to men^19,20^. In addition, sex hormones may differentially impact the frequency and severity of many autoimmune and inflammatory diseases, generally more prevalent in women than men^21,22^. It should be noted that sex hormones may exert apparently contrasting effects. For example, an immunosuppressive role for testosterone was observed in response to influenza vaccination^23^, while testosterone supplementation following influenza infection in aged male mice, which caused decreased serum testosterone levels, reduced mortality^24^.

In order to better understand the relationship between testosterone status and disease severity, we have studied a population of male and female COVID-19 patients for serum and blood biomarkers in association with disease outcome, and have performed a longitudinal analysis in a sub-cohort of male patients. We have found that male patients who survive the disease eventually reinstate physiological levels of testosterone, while those who die from COVID-19 fail to do so. As such, we show that the trajectories of serum testosterone levels are highly accurate predictors of survival [area under the curve (AUC) of receiver operating characteristic (ROC) curve = 92.8%, *p* < 0.0001] in male COVID-19 patients, independent of associated co-morbidities or clinical management, and superior in predictive power to bloodlymphocyte or neutrophil counts. Furthermore, we have found that male COVID-19 patients with a fatal outcome display a late coordinated depletion of circulating subsets of differentiated CD4+ T lymphocytes and monocytes, mirrored by a relative enrichment of undifferentiated CD4+ T cells and monocytes.

## Results

With the aim of exploring factors that may underlie the worse progression of COVID-19 in men, we have undertaken a retrospective study of 249 male and 248 female patients admitted to the Vall d’Hebron Hospital between May1^st^ and June 30^th^ 2020, with RT-PCR-confirmed diagnosis of SARS-CoV-2 infection. Patients were first studied for serum biochemical and hematological variables in samples collected at or near admission (first time point or Sample 1), for baseline assessment. Subsequently, a selected male population (114 patients) was studied for the progression of the disease by analyzing the same variables longitudinally, with serial time-point sample collection on average every 7 days. Samples from this patient subcohort were further analyzed for serum luteinizing hormone (LH) and androstenedione levels, as well as an extensive phenotyping of circulating immune cells.

### Biochemical and hematological predictors of outcome in male and female COVID-19 patients

Biochemical and hematological parameters were analyzed at admission for 497 male and female patients. Demographics, background information and treatments are shown in Tables 1 and 2 and Fig. SF1 and Tables ST1 and ST2. Patients were grouped according to their eventual outcomes at discharge/death into mild, moderate, severe-survivor and severe-deceased (Table ST3). In spite of the comparable overall age distributions in male and female patients (Fig. SF1), a higher proportion of female patients fell into the mild and moderate outcome groups (56%) as compared to males (44%). Conversely, a higher proportion of male patients fell into the severe-survivor and severe-deceased outcome groups (56% and 58.46%, respectively) as compared to female patients (44% and 41.5%). In both male and female patients, the median age severe-deceased outcome groups were significantly older than those in mild or moderate outcome groups, as expected^25^. However, women in the severe-deceased group were significantly older than males in the same outcome group (Fig. SF1).

**Table 1.** Baseline clinical characteristics of the male study population.

|  | **Mild-Moderate**<br>**N=114** | **Severe-recovered**<br>**N=97** | **Severe-deceased**<br>**N=38** | **p-value<sup>1-2</sup>** | **p-value<sup>1-3</sup>** | **p-value<sup>2-3</sup>** |
| --- | --- | --- | --- | --- | --- | --- |
| *Age (y), median (IQR)* | 59 (56-63) | 56 (53-59) | 68 (67-71) | 0.1096 | 0.0001 | <0.0001 |
| *Length of stay (days), median (IQR)* | 8 (7-9) | 30.5 (27-34) | 19 (9-26) | <0.0001 | <0.0001 | 0.0009 |
| *Comorbidity, n° (%)* | 54 (47.36) | 57 (58.76) | 28 (73.68) | 0.1279 | 0.0051 | 0.1175 |
| *Hypertension* | 35 (30.70) | 31 (31.96) | 21 (55.26) | 0.8822 | 0.0109 | 0.0178 |
| *Diabetes* | 20 (17.54) | 15 (16.46) | 11 (28.95) | 0.7143 | 0.1630 | 0.0908 |
| *Cancer* | 6 (5.26) | 3 (3.09) | 3 (7.89) | 0.5115 | 0.6915 | 0.3497 |
| *Cardiovascular diseases* | 9 (7.89) | 12 (12.37) | 3 (7.89) | 0.3573 | 0.9999 | 0.5560 |
| *Chronic Lung diseases* | 10 (8.77) | 6 (6.19) | 5 (13.16) | 0.6046 | 0.5297 | 0.2912 |
| *Obesity-Dyslipidaemia* | 16 (14.03) | 33 (34.02) | 11 (28.95) | 0.0009 | 0.0497 | 0.6842 |
| *Chronic Kidney disease* | 6 (5.26) | 2 (2.06) | 1 (2.63) | 0.2927 | 0.6808 | 0.6727 |
| *Others* | 9 (7.89) | 13 (13.4) | 7 (18.42) | 0.2586 | 0.1220 | 0.5903 |
| *Biochemical parameters, median (IQR)* |  |  |  |  |  |  |
| *Leucocytes n°* | 6.32 (5.78-6.88) | 7.80 (6.83-8.80) | 8.70 (7.20-9.65) | <0.0001 | 0.0051 | 0.9999 |
| *Lymphocytes n°* | 1.20 (1.0 -1.4) | 0.77 (0.70- 0.82) | 0.8(0.65-0.90) | <0.0001 | 0.0001 | 0.9999 |
| *Lymphocytes %* | 18.55(16.15– 1.57) | 9.20 (8.18- 11.28) | 10.89 (7.95 -13.38) | <0.0001 | <0.0001 | 0.9999 |
| *Neutrophils* | 4.57 (3.92-4.90) | 6.26 (5.53 – 7.38) | 6.39 (5.53-8.34) | <0.0001 | 0.0007 | 0.9999 |
| *Interleukine-6* | 42.30(25.33-53.46) | 130.8(112.10-160.6) | 215.30(117.6-996.4) | <0.0001 | <0.0001 | 0.9999 |
| *C-reactive protein* | 9.57 (6.76-10.88) | 17.47(12.17-0.72) | 14.56(9.84 – 24.019) | <0.0001 | 0.0069 | 0.9999 |
| *LDH* | 324 (296.0-369.0) | 472.5(440.0-506.0) | 493.0(429.0-583.0) | <0.0001 | <0.0001 | 0.9999 |
| *D-Dimer* | 236 (213.0 -311.0) | 529.0 (361.0 -711.0) | 421.5 (260.0-684.0) | <0.0001 | 0.0210 | 0.9999 |
| *Ferritin* | 934 (679.0 -1089.0) | 1289 (1100.0-1574.0) | 1443.0(554.9-1919.0) | 0.0022 | 0.7403 | 0.6783 |

**Table 2.**
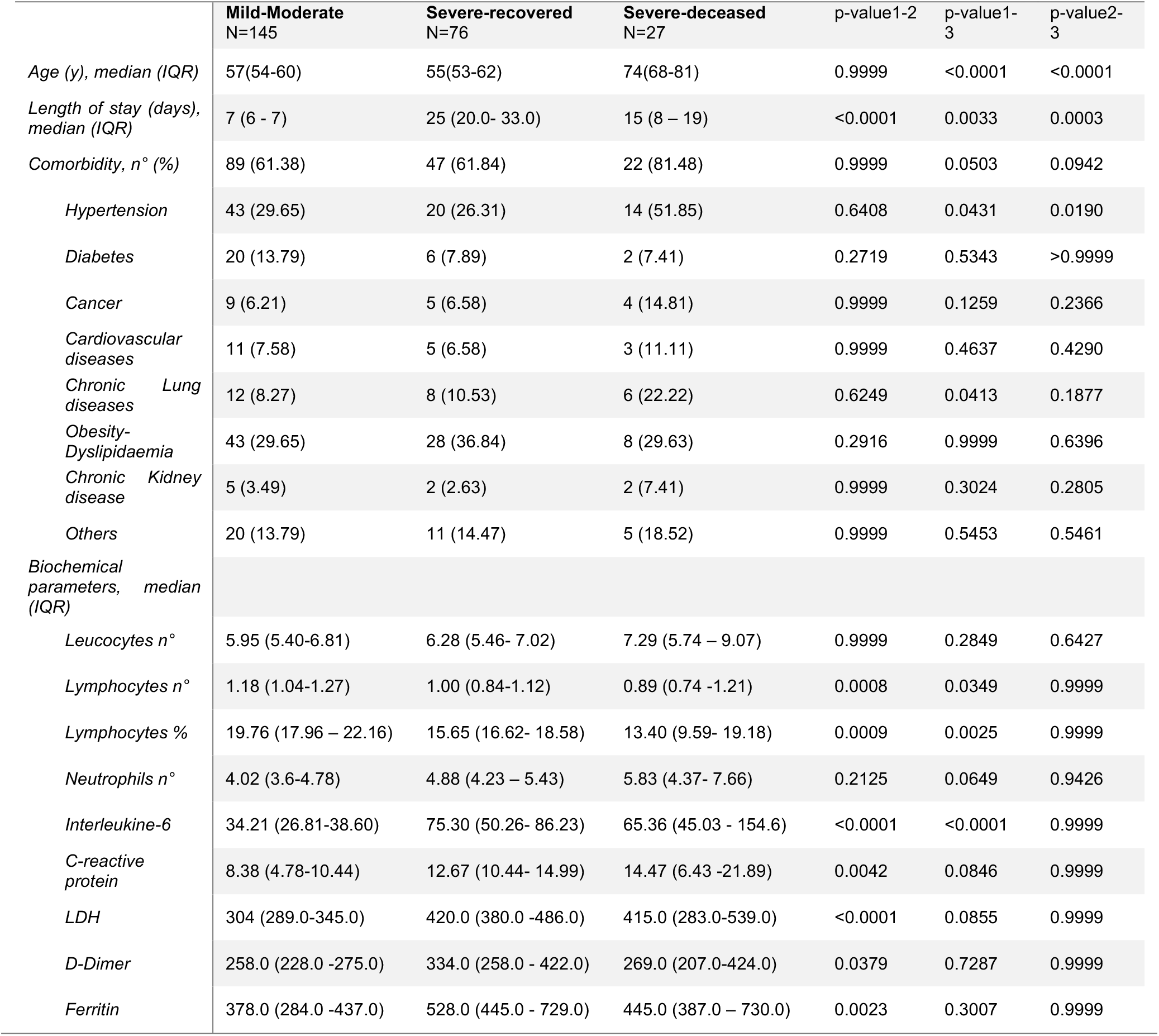
Baseline clinical characteristics of the female study population.

|  | **Mild-Moderate**<br>**N=145** | **Severe-recovered**<br>**N=76** | **Severe-deceased**<br>**N=27** | **p-value1-2** | **p-value1-3** | **p-value2-3** |
| --- | --- | --- | --- | --- | --- | --- |
| *Age (y), median (IQR)* | 57(54-60) | 55(53-62) | 74(68-81) | 0.9999 | <0.0001 | <0.0001 |
| *Length of stay (days), median (IQR)* | 7 (6 - 7) | 25 (20.0- 33.0) | 15 (8 – 19) | <0.0001 | 0.0033 | 0.0003 |
| *Comorbidity, n° (%)* | 89 (61.38) | 47 (61.84) | 22 (81.48) | 0.9999 | 0.0503 | 0.0942 |
| *Hypertension* | 43 (29.65) | 20 (26.31) | 14 (51.85) | 0.6408 | 0.0431 | 0.0190 |
| *Diabetes* | 20 (13.79) | 6 (7.89) | 2 (7.41) | 0.2719 | 0.5343 | >0.9999 |
| *Cancer* | 9 (6.21) | 5 (6.58) | 4 (14.81) | 0.9999 | 0.1259 | 0.2366 |
| *Cardiovascular diseases* | 11 (7.58) | 5 (6.58) | 3 (11.11) | 0.9999 | 0.4637 | 0.4290 |
| *Chronic Lung diseases* | 12 (8.27) | 8 (10.53) | 6 (22.22) | 0.6249 | 0.0413 | 0.1877 |
| *Obesity-Dyslipidaemia* | 43 (29.65) | 28 (36.84) | 8 (29.63) | 0.2916 | 0.9999 | 0.6396 |
| *Chronic Kidney disease* | 5 (3.49) | 2 (2.63) | 2 (7.41) | 0.9999 | 0.3024 | 0.2805 |
| *Others* | 20 (13.79) | 11 (14.47) | 5 (18.52) | 0.9999 | 0.5453 | 0.5461 |
| *Biochemical parameters, median (IQR)* |  |  |  |  |  |  |
| *Leucocytes n°* | 5.95 (5.40-6.81) | 6.28 (5.46- 7.02) | 7.29 (5.74 – 9.07) | 0.9999 | 0.2849 | 0.6427 |
| *Lymphocytes n°* | 1.18 (1.04-1.27) | 1.00 (0.84-1.12) | 0.89 (0.74 -1.21) | 0.0008 | 0.0349 | 0.9999 |
| *Lymphocytes %* | 19.76 (17.96 – 22.16) | 15.65 (16.62- 18.58) | 13.40 (9.59- 19.18) | 0.0009 | 0.0025 | 0.9999 |
| *Neutrophils n°* | 4.02 (3.6-4.78) | 4.88 (4.23 – 5.43) | 5.83 (4.37- 7.66) | 0.2125 | 0.0649 | 0.9426 |
| *Interleukine-6* | 34.21 (26.81-38.60) | 75.30 (50.26- 86.23) | 65.36 (45.03 - 154.6) | <0.0001 | <0.0001 | 0.9999 |
| *C-reactive protein* | 8.38 (4.78-10.44) | 12.67 (10.44- 14.99) | 14.47 (6.43 -21.89) | 0.0042 | 0.0846 | 0.9999 |
| *LDH* | 304 (289.0-345.0) | 420.0 (380.0 -486.0) | 415.0 (283.0-539.0) | <0.0001 | 0.0855 | 0.9999 |
| *D-Dimer* | 258.0 (228.0 -275.0) | 334.0 (258.0 - 422.0) | 269.0 (207.0-424.0) | 0.0379 | 0.7287 | 0.9999 |
| *Ferritin* | 378.0 (284.0 -437.0) | 528.0 (445.0 - 729.0) | 445.0 (387.0 – 730.0) | 0.0023 | 0.3007 | 0.9999 |

As an approach to capture global patterns of association between biochemical parameters and outcomes, we applied principal component analysis (PCA), followed by Spearman multivariate correlation analysis (Fig. 1a, b). In male patients (Fig. 1a), we observed a clear correlation between mild or moderate outcomes with known predictors of good outcome, such as higher lymphocyte counts or hemoglobin levels, while severe outcome groups correlated with low neutrophil counts, and high IL-6, CRP, D-dimer, ferritin or LDH levels, confirming abundant prior evidence^1,25,26^. Patients with moderate outcomes had significantly lower serum testosterone levels compared to those with mild outcomes (Fig. 1c), in agreement with other studies^12,13^. Additionally, severe outcome patients had significantly lower serum testosterone levels compared to those with mild or moderate outcomes (Fig. 1c). Interestingly, the low testosterone levels found in the admission time-point determinations for both severe outcome groups were not significantly different between survivor and deceased patients (Fig. 1c). Furthermore, older age presented a stronger correlation with a severe-deceased outcome than biochemical parameters predictive of poor outcome, such as D-dimer, ferritin, LDH or IL-6 (Fig. 1a), whereas the reverse was the case for correlations with a severe-survivor outcome, in which biochemical parameters were more strongly correlated with this group than age (Fig. 1a).

**Figure 1.**
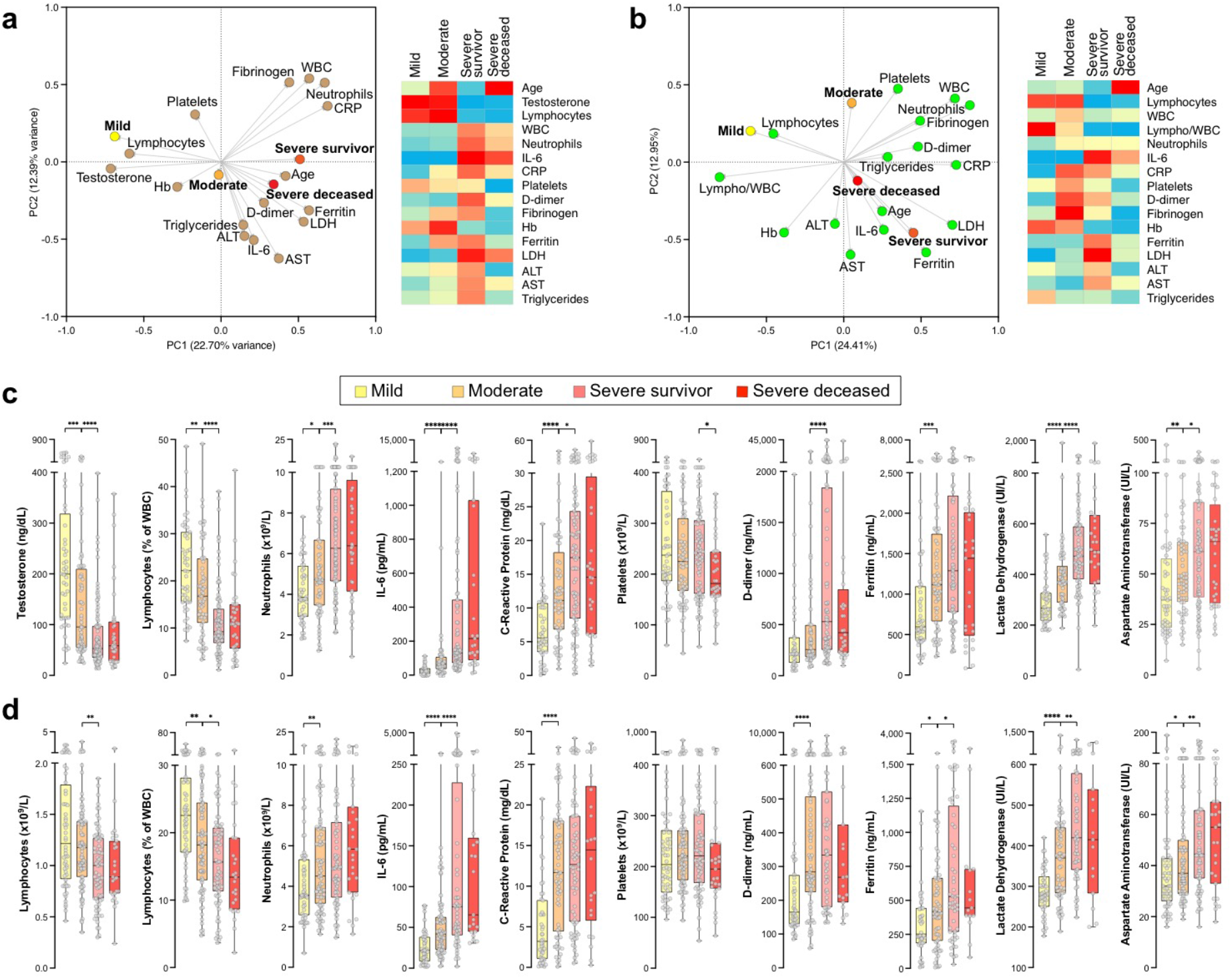
Clinical biochemistry features of male (**a, c**) and female (**b, d**) COVID-19 patients, associated with outcomes. Clinical biochemistry values were determined for samples collected at patient admission. **a, b**. <u>Left panels:</u> Principal component analysis (PCA) illustrating correlations between elevated levels of the indicated parameters and mild, moderate, severe survivor or severe deceased outcomes in male (**a**) or female (**b**) patients. <u>Right panels:</u> Heatmap of correlation coefficients between elevated levels of biochemical parameters and outcomes. Spearman multivariant correlation analyses were performed for all parameters *vs*. outcomes, the resulting coefficients normalized for each column (range, 0 to 1) and used to build heatmaps. **c, d**. Values of relevant clinical biochemistry parameters assessed for admission samples, and grouped by eventual outcome for male (**c**) and female (**d**) patients. Asterisks denote significance of pairwise comparisons between samples grouped by outcome, determined by t-test analysis: *p ≤ 0.05; **p ≤ 0.01; ***p ≤ 0.001; ****p ≤ 0.0001. Non-significant comparisons (p > 0.05) are not shown.

In female patients, PCA and multivariate analysis also highlighted significant differences between mild-moderate and severe outcome groups (Fig. 1b and d). Similar to male patients, the mild outcome group of female patients showed strong correlations to lymphocyte counts and hemoglobin levels, while the severe outcome groups were correlated with high IL-6, CRP, D-dimer, ferritin and LDH, (Fig. 1b and d). Mirroring male patients, older age showed the strongest correlation to a severe-deceased group in female patients (Fig. 1b). Age, platelet counts and fibrinogen levels significantly discriminated severe-survivor from severe-deceased female patients (Fig. 1 b and d).

The risk of ICU admission for patients with mild-moderate outcomes was assessed by odds ratio (OR) estimates and logistic regression analysis. In male patients, the most significant OR of ICU admission were found for IL-6 (OR 12.94, 95% CI 7.73-24.72), LDH (OR 7.88, 95% CI 4.81-14.17), lymphopenia (OR 0.15, 95% CI 0.09-0.25) and neutrophilia (OR 4.02, 95% CI 2.53-6.71) (Fig. 2a), in agreement with previous studies^25,27^. A significant OR was also found for testosterone (0.16, 95% CI 0.10-0.26) also in line with other studies^12,13^. In female patients, the most significant OR of ICU admission were for IL-6 (OR 6.41, 95% CI 3.86-11.60), LDH (OR 4.86, 95% CI 2.82-9.15), aspartate aminotransferase (OR 2.58, 95% CI 1.60-4.28) and lymphocytes percentage of WBC (OR 0.34, 95% CI 0.21-0.54) (Fig. 2a). Interestingly, testosterone levels at admission were not significantly associated with the occurrence of comorbidities in older men (Fig. SF2).

**Figure 2.**
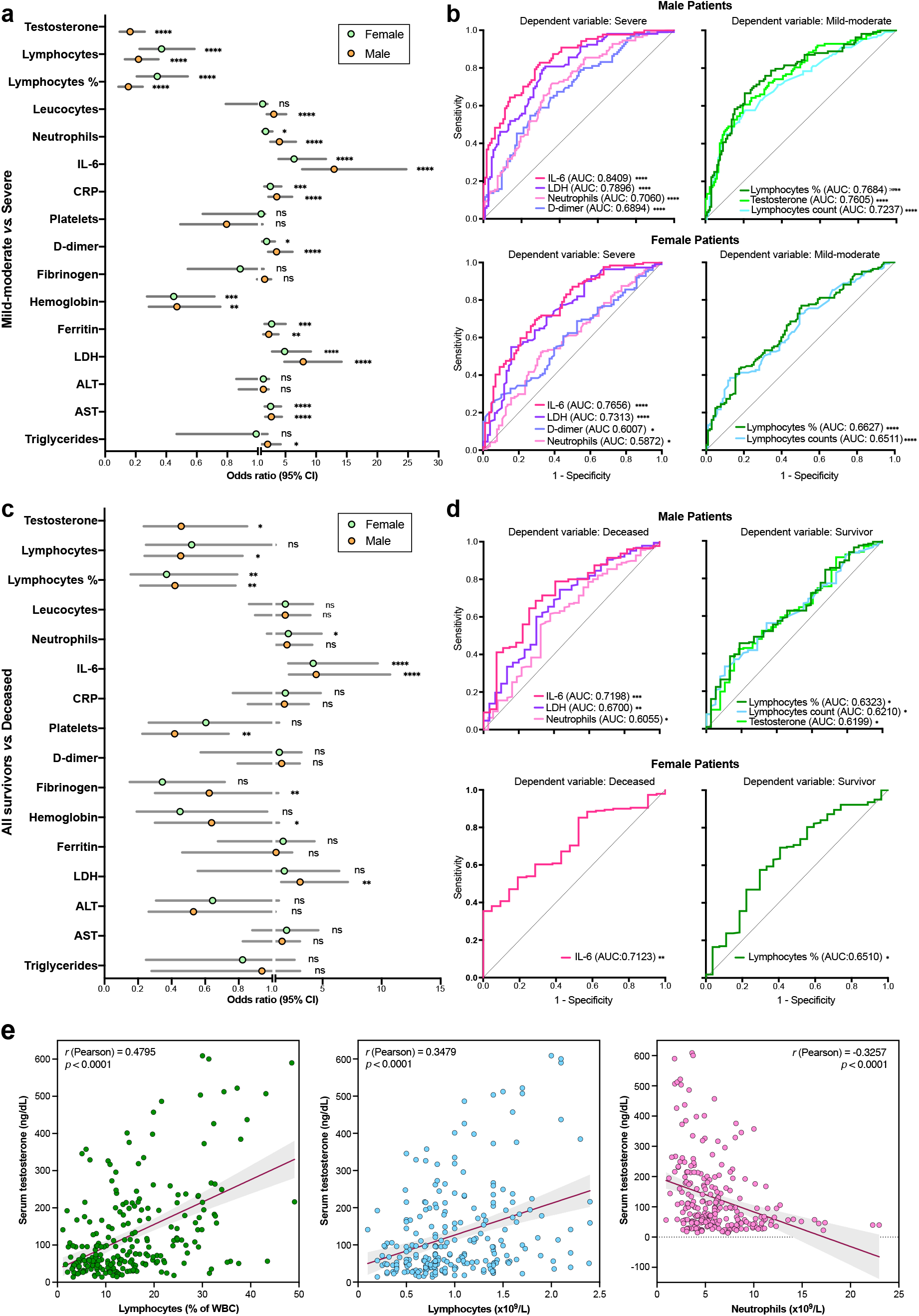
Assessment of clinical biochemistry parameters as predictors of risk of severe disease or death from COVID-19. **a, c**. Odds ratios (OR) of clinical biochemistry parameters and risk of severe disease (**a**) or death (**c**) in male and female patients. **b, d**. Receiver operating characteristic (ROC) curves and area under the curve (AUC) values of risk of severe disease (**b**) or death (**d**). Shown are only those parameters with significant AUC values (p ≤ 0.05). **e**. Correlations of testosterone serum levels with lymphocytes (percentage of WBC and counts), and neutrophil counts.

The power of these parameters to predict severe disease was corroborated by multiple logistic regression analysis. The resulting receiver operating characteristic (ROC) curves yielded areas under the curve (AUC), which, in male patients, were > 0.7 (*p* < 0.0001) for IL-6, LDH and neutrophilia, and < 0.23 for lymphopenia and testosterone. In females, AUC of ROC curves were > 0.7 (*p* < 0.0001) for IL-6 and LDH (Fig. 2b).

The same parameters showed a weaker power to predict risk of death from COVID-19, with the exception of serum IL-6 levels, in both male and female patients (OR 4.45, 95% CI 2.14 – 10.71 and AUC of ROC 0.7189, *p* = 0.0002 for males; and females had OR 4.21 and AUC of ROC 0.7123, *p* = 0.0014 for females) (Fig. 2c, d).

In sum, this baseline analysis reveals that critically low serum testosterone levels in male patients are a risk factor for severe COVID-19. Although other factors predictive of severity are in line with prior evidence^25,27^, our observations also suggest that, compared to age-matched female patients, male COVID-19 patients present a higher risk of progression to a severe-critical disease, with higher levels of inflammatory markers (serum IL-6, blood neutrophil counts) and tissue damage (LDH), and more marked lymphopenia^2^. Importantly, biochemical and hematological parameters determined at or near hospital admission do not reveal strong predictive markers of death from COVID-19, with the exception of serum IL-6 levels. A significant correlation of serum testosterone levels with lymphocyte (absolute counts, r = 0.3122; percentages of WBC, r = 0.4187) and neutrophil (r = -0.3586) counts suggests that these three parameters may be mutually coupled (Fig. 2e).

### Recovery of serum testosterone levels accurately predicts survival in male COVID-19 patients

In order to explore whether determinations in longitudinal samples later in the clinical course could yield additional or improved predictors of lethal disease, we followed the progression of a sub-cohort of 114 male COVID-19 patients, by analyzing samples procured at different time points after hospital admission. For patients in the severe outcome group, up to five time-point samples were analyzed. Patients in the combined mild-moderate outcome group were discharged, in average, after two weeks of admission and a maximum of three time-point samples were procured from them.

Trajectories for all clinical biochemistry and hematological parameters under consideration were plotted for all patients grouped into mild-moderate, severe survivor and severe deceased outcomes (Fig. 3a and Fig. SF3). Average values for each parameter were calculated for each time-point and linear regression applied. The trajectories of only three parameters, namely testosterone (*p* = 0.0038), lymphocyte counts (or percentage of WBC) (*p* = 0.01) and neutrophil counts (*p* = 0.0023), were significantly different, as analyzed by two-way ANOVA, in comparisons of trajectories between the severe survivor and severe deceased groups (Fig. 3a). None of the other parameters analyzed showed statistically significant divergent trajectories in comparisons between severe survivor *vs*. severe deceased outcomes (Fig. SF3).

**Figure 3.**
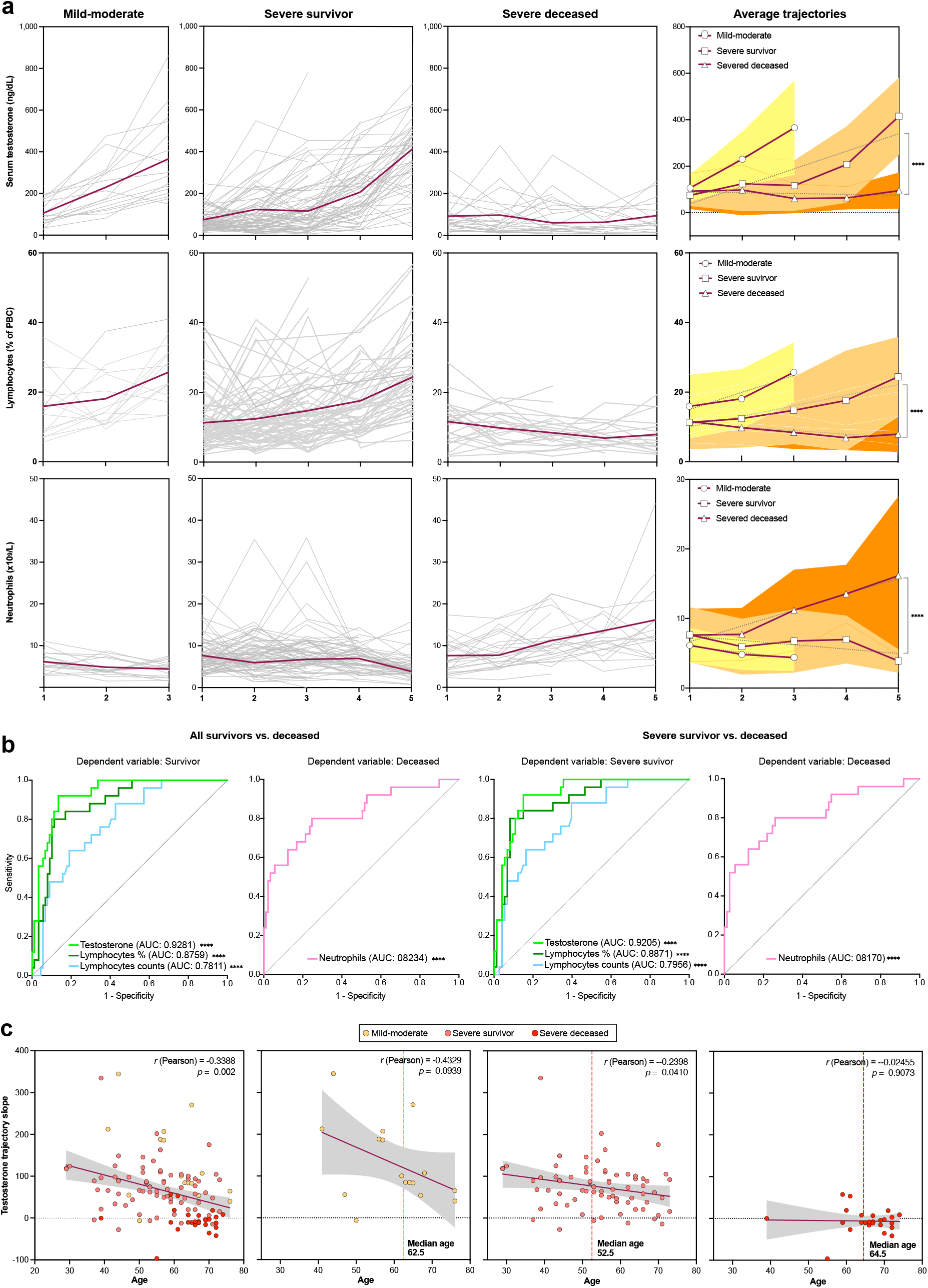
Recovery of serum testosterone levels and blood lymphocyte counts predict survival in male COVID-19 patients. **a**. Longitudinal determinations (≥ 3 samples per patient collected on separate dates) of clinical biochemistry parameters were performed, and trajectories for individual patients (grey lines) and average values (red lines) plotted. A given time-point corresponds to a cluster of days post-admission (± 3 days). Linear regression was applied to average trajectories and the resulting slopes compared for significance between outcome groups by means of two-way ANOVA. **b**. ROC curves and AUC values for longitudinal trajectories (linear regression slopes) of serum testosterone, blood lymphocyte counts (number per mL and % of white blood cells) and blood neutrophils as predictors of survival in comparisons of all surviving *vs*. deceased patients (<u>left two panels</u>) or surviving patients with severe disease *vs*. deceased patients (<u>right two panels</u>). Longitudinal analyses for additional clinical biochemistry parameters are shown in Extended Data Fig. 3. **c**. Correlations of age with testosterone trajectory slopes in all patients with longitudinal analyses (leftmost panel) and in different outcome groups.

The values of the slopes calculated from the linear regression of the trajectories of all parameters and patients were used in multiple logistic regression analysis to estimate their outcome predictive power. The resulting ROC curves and AUC values indicated that serum testosterone trajectories are remarkably accurate predictors of survival from COVID-19, both in all-survivor *vs*. deceased (AUC = 0.9281, 95% CI 0.7216 to 0.9252, *p* < 0.0001) and severe survivor *vs*. deceased outcome (AUC = 0.9205, 95% CI 0.8664 to 0.9747, *p* < 0.0001) comparisons (Fig. 3b). Lymphocyte counts (absolute numbers per dL or percentage of WBCs) were also highly accurate predictors of outcome, as were neutrophil counts (Fig. 3b). Interestingly, the trajectories of IL-6 or LDH, which values on admission were predictive of severity and death from COVID-19 in male patients, were not significantly different in these longitudinal comparisons (Fig. SF3).

Age is a factor predictor of COVID-19 severity^25^. In our cohort of male patients, the testosterone trajectory slopes significantly and inversely correlated with age (*r* = -0.3801, p < 0.0001) (Fig. 3c). Consistently, a majority of patients with severe deceased outcomes had low or negative testosterone trajectory slopes (Fig. 3c). However, and interestingly, the median age of patients with severe deceased outcomes was not significantly different from the median age of patients with mild-moderate outcomes (Fig. 3c), and a substantial proportion of patients with severe survivor outcomes were aged older than 60 (Fig. 3c). Likewise, while the frequency of comorbidities was higher among patients with fatal outcomes as compared with those who survived severe disease, it was not significantly different from the frequency of the moderate outcome group (Fig. SF1). This suggests that old age, with or without accompanying comorbidities, may impact the ability of a subset of COVID-19 patients to reinstate testosterone production, coupled to a failure to recover from the disease.

These results suggest a role for testosterone in deregulation of the immune response in deceased patients, in particular for the observed lymphopenia and neutrophilia.

### The LH-androstenedione axis is not significantly perturbed in male COVID-19 patients

Testosterone is largely synthesized from cholesterol through androstenedione in the Leydig cells of the testis under the stimulus of luteinizing hormone (LH), secreted from the anterior portion of the pituitary gland^28^. The observed critical decline in circulating testosterone levels in male COVID-19 patients suggests the occurrence of a transient (survivor outcomes) or sustained (fatal outcomes) hypogonadism following the onset of COVID-19, for which several causal mechanisms may be proposed. One possible mechanism is an inhibition of the LH-androstenedione axis, previously associated with non-specific critical illness^29^ and the deleterious action on the hypophysis of inflammatory cytokines^30^ . A second possible mechanism may involve infection and damage by SARS-CoV-2 of testicular cells expressing ACE2, mainly Leydig cells^31,32^. In the first scenario, acute declines in LH and adrostenedione levels would be expected, while in the latter scenario they would be either unaffected or increased for LH due to a negative feedback loop with testosterone^29^.

We determined circulating LH and androstenedione levels in a longitudinal series of samples in a patient subcohort for which testosterone trajectories had been concomitantly determined, as described above. The median levels of LH were within normal ranges, independent of patient outcome (Fig. 4a). Similarly, longitudinal LH trajectories were not significantly different between patients in the survivor *vs*. deceased outcomes, in stark contrast with the strongly divergent testosterone trajectories (Fig. 4b). Nevertheless, LH levels determined in the last of the longitudinal samples showed a decline in the deceased outcome group as compared to the severe survivor group, although without reaching statistical significance. On the other hand, although androstenedione levels fell within normal ranges in the majority of patients in all outcome groups and throughout the longitudinal analysis (Fig. 4a, b), deceased patients showed an increase as compared to the survivor outcome groups, without a concomitant increase in testosterone levels (Fig. 4a). It should be noted that circulating androstenedione is produced mostly by adrenal glands^28^ and its synthesis might be affected by the corticosteroids used to treat these patients that may block the endogenous production of cortisol, corticosterone and aldosterone, deviating the synthesis from cholesterol to androstenedione^28^.

**Figure 4.**
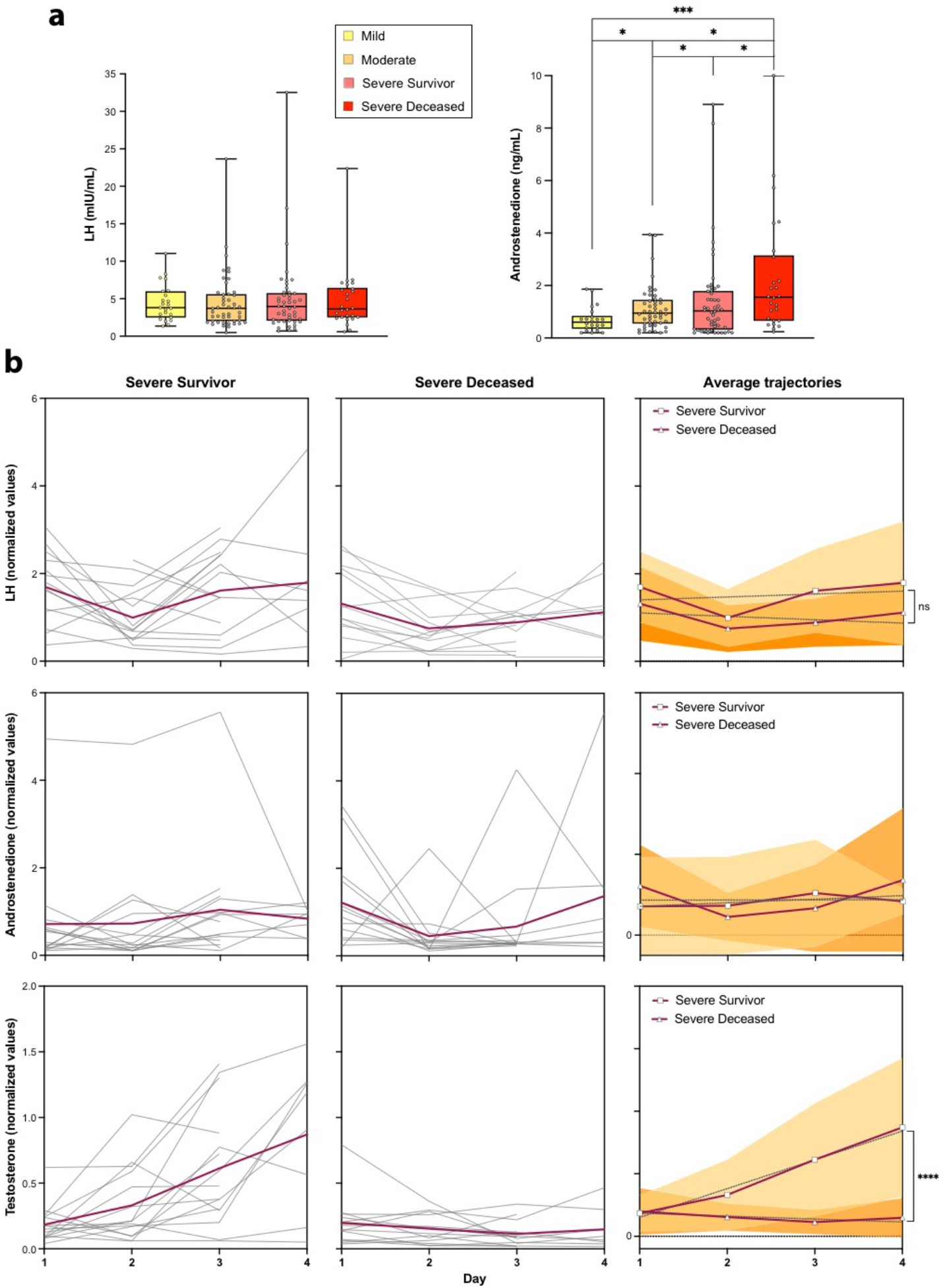
The luteinizing hormone (LH)-androstenedione axis is not significantly perturbed in male COVID-19 patients. **a**. Determinations of serum LH and androstenedione levels in samples collected at admission, grouped by eventual outcomes. Pair-way between-group comparisons were performed by t-test. **b**. Longitudinal determinations (≥ 3 samples) of serum LH, androstenedione and testosterone levels, analysed as in Fig. 3. Comparisons of trajectories (linear regression slopes) were performed by two-way ANOVA.

Therefore, the failure of patients with fatal outcomes to recover their physiological levels of testosterone in spite of LH and androstenedione levels within normal ranges suggests the development of an irreversible peripheral failure in the biosynthesis of testosterone in these patients. As such, an irreversible damage of Leydig cells^33^ in patients with fatal outcomes could explain these observations, while a resolution of viral infection would explain the recovery of a normal production of testosterone in survivors.

### Lethal male COVID-19 is associated with a depletion of circulating T helper cells

A number of studies have found substantial differences in immune responses to SARS-CoV-2 between male and female patients^15,17^, although the mechanisms underlying these differences are still unclear. In order to address the relationship between testosterone trajectories, outcome and immune status in male patients, we analyzed circulating immune subpopulation repertoires in a subset of our patient cohort, in at least two independent determinations, separated by 5-20 days.

Principal component analyses (PCA) pointed to a correlation between immune subpopulations, outcome and testosterone levels both in a first determination of samples near admission date (Sample 1) and a subsequent analysis of samples near discharge or death date (Sample 2), with some subpopulations showing remarkable shifts between Sample 1 and Sample 2 in their correlations with outcome (Fig. 5a, b). As such, Sample 1 determinations demonstrated relatively few changes in immune cell repertoires between surviving and deceased patients. In stark contrast, a subsequent determination (Sample 2) showed a coordinated depletion of T helper subpopulations in association with death, along with changes in natural killer (CD56^+^brightCD16^-^ and CD56^+^dimCD16^+^) and monocyte subpopulations (Fig. 5a). The subpopulations with the most significant changes in relative abundance as a function of outcome tended to correlate with serum testosterone levels sampled in the same period (1-3 days from sampling for immune repertoire analyses) (Fig. 5a).

**Figure 5.**
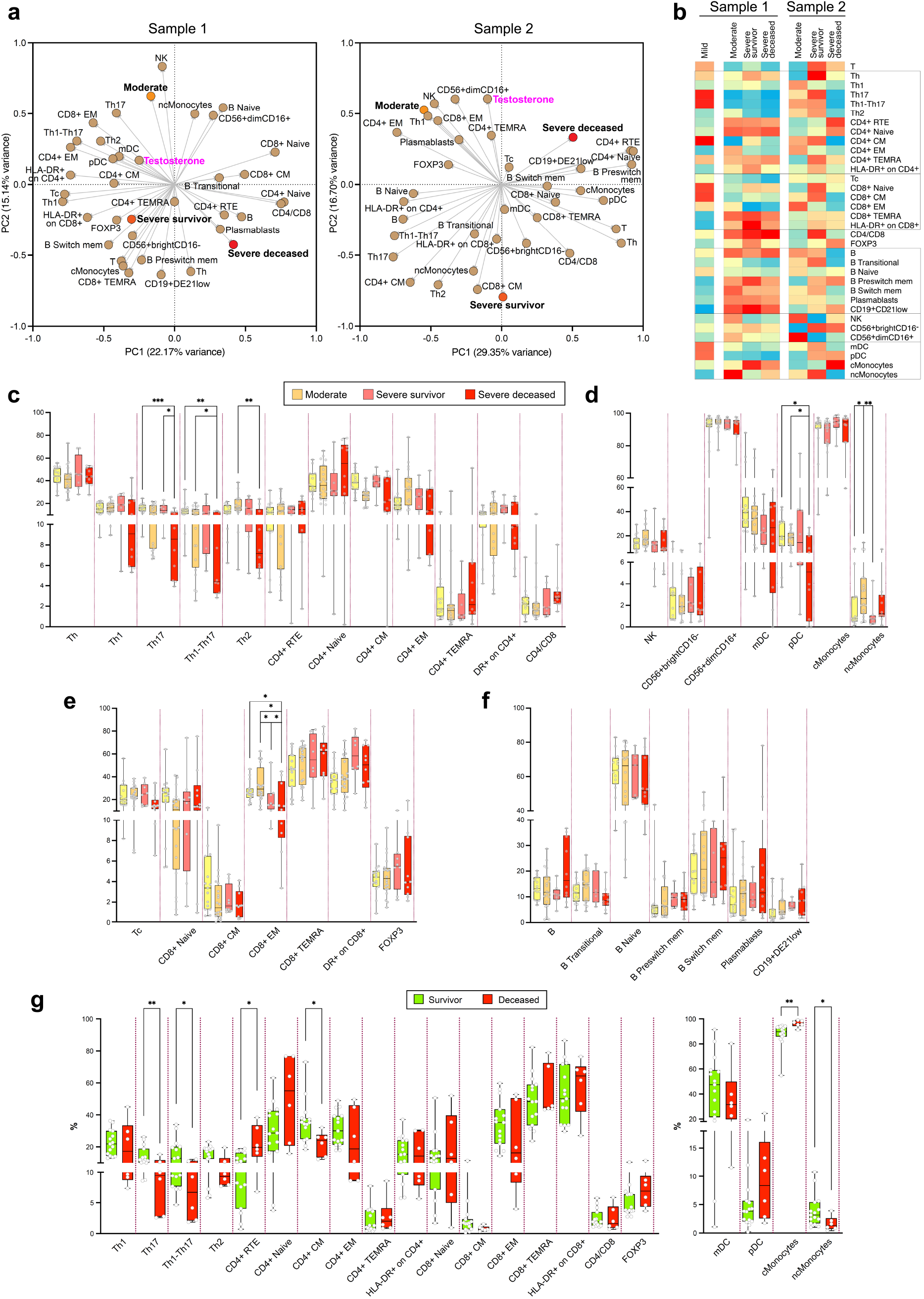
Immune switch during the course of disease in severe and deceased patients, as determined by multiparameter profiling of circulating immune cells. **a**. PCA of samples analysed near admission (<u>Sample 1</u>) and near discharge or death (<u>Sample 2</u>). Mild, moderate, severe survivor and severe deceased outcomes were assigned values 1, 2, 3 and 4, respectively. Serum testosterone values of samples collected in the same or nearby dates (± 3 days) were included in the analysis. The indicated immune subpopulations are defined by cell-surface markers and determined by spectral flow cytometry (Materials and Methods). **b**. Heatmap of correlation coefficients between immune subpopulation values and outcomes, for near-admission (<u>Sample 1</u>) and near-discharge/death (<u>Sample 2</u>) samples. Spearman multivariant correlation analyses were performed for all parameters *vs*. outcomes, the resulting coefficients normalized for each column (range, 0 to 1) and used to build heatmaps. **c-f**. Between-outcome comparisons of immune cell subpopulation repertoires: CD4+ (**c**); natural killer, dendritic and monocyte (**d**); CD8+ (**e**); and B (**f**) cell subpopulations. **g**. Survivor (mild, moderate, severe survivor) *vs*. deceased patient comparisons for T cell (CD4+ and CD8+) and dendritic cells and monocytes. Such comparisons were not significant for other immune subpopulations (B cells, NK cells).

These correlations were more evident in multivariate correlation analyses, particularly for Sample 2 (Fig. 5b). These analyses show a generalized loss of representation of circulating CD4^+^ (CD4^+^ central memory, CD4^+^ effector memory, CD4^+^ TEMRA) and T helper (Th) T lymphocyte subpopulations (Th1, Th17, Th1-Th17, Th2) in deceased patients compared to surviving patients, accompanied with a reciprocal increased representation of naïve CD4^+^ T cell compartments (recent thymus emigrant (RTE) CD4^+^, naïve CD4^+^CD45RA^+^CCR7^+^) (Fig. 5b-d). They also point to an association of monocyte differentiation with outcome, with a predominant correlation with non-classical monocytes in moderate and severe survivor patients and, conversely, with more pro-inflammatory^34^ classical monocytes in deceased patients (Fig. 5b-d). There are additional associations of cytotoxic T cell (particularly diminished CD8^+^ effector memory) or B cell subpopulations (diminished transitional B cells and augmented representation of plasmabasts) with lethal COVID-19 (Fig. 5 d-f), albeit without reaching statistical significance. These correlations are made more evident when comparing all surviving patients (moderate and severe survivor) with deceased patients (Fig. 5g), and further illustrated by flow cytometry histograms of representative cases (Fig. SF4).

Together, these observations indicate that male COVID-19 patients with a lethal outcome suffer from a decline in circulating effector T helper cells accompanied with a relative enrichment in circulating naïve CD4^+^ T cells. These selective imbalances in immune subpopulations in association with fatal outcomes are in line with other studies^34-37^ and suggest a defective transition from undifferentiated to differentiated T helper cells^38^ and classical to non-classical monocytes^34,35^, an altered leucocyte recirculation dynamics with effect on naïve/effector T lymphocyte balance^36^, or exhaustion^39,40^ of differentiated effector T helper cells and non-classical monocytes. Failure to recover from such imbalances is paralleled by a failure to reinstate physiological levels of testosterone and is associated with a lethal outcome in male patients.

## Discussion

Numerous studies have identified prognostic markers able to discern severe COVID-19 patients, including older age, male sex, co-morbidities such as obesity, diabetes or cardiovascular disease, elevated circulating markers of inflammation, lymphopenia, neutrophilila^2,12,16,25-27,41^, or the presence of autoantibodies to class-I interferons^42^. Nomograms or scores that combine several independent parameters have been proposed as predictors of COVID-19 outcome^2,12,25^. However, relatively few studies have addressed sex differences in predictive markers of disease outcome^2,12,25,43-45^. Interestingly, inflammation markers, but not co-morbidities, BMI or age have been found to be associated with outcome differences between male and female COVID-19 patients^45^

Our comparative analysis of biochemical and hematological parameters has revealed that both sexes share markers with significant predictive power of disease outcome, including IL-6, LDH, D-dimer, lymphopenia and neutrophilia, although the levels of these markers, and the strength of their predictive power, are consistently higher in male patients as compared to female patients. This becomes more evident when evaluating predictive markers of lethal COVID-19, which yields IL-6 and lymphocyte (percentage of total WBC) as the only two significantly predictive markers shared in both male and female patients. Other significant markers predictive of lethal COVID-19 in male, but not female, patients are LDH levels, neutrophilia and absolute lymphocyte counts, in addition to testosterone levels, which are exclusively masculine in our patient cohorts. These observations suggest that male COVID-19 patients with severe and lethal disease suffer from more deleterious underlying pathogenic and inflammatory processes than female patients with comparable clinical severity^2^, a situation also observed in other respiratory viral infections^4,5^.

Interestingly, markers of inflammation (IL-6, CRP) or tissue damage (LDH), with good outcome predictive power in first sample determinations, lost their predictive power in longitudinal analyses of samples in male patients, collected up to the time of discharge or fatality. In contrast, testosterone levels, whose determinations on admission provided relatively modest outcome predictive power, corroborating other studies^11,13^, gained remarkable levels of significance when analyzed longitudinally. The AUC values of ROC curves in multiple logistic regression analyses (mild-moderate *vs*. severe: 0.9281, 95% CI 0.7216 to 0.9252, *p* < 0.0001; severe survivor *vs*. deceased: 0.9205, 95% CI 0.8664 to 0.9747, *p* < 0.0001) indicate that serum testosterone trajectories in longitudinal determinations constitute, to the best of our knowledge, the most accurate independent predictors of disease outcome in male COVID-19 patients described so far. Furthermore, longitudinal trajectories of lymphocyte and neutrophil counts also yield highly significant predictions of disease outcome.

Other biochemical parameters indicative of pathological inflammatory or pro-coagulant states, such as elevated IL-6, CRP or D-dimer levels, eventually return to near-physiological levels in both survivors and patients with fatal outcomes. This has been observed in other studies^45,46^, and suggests that the normalization of these factors is insufficient, *per se*, to abate the pathological hyperinflammation and hypercoagulation accompanying severe COVID-19 with a fatal outcome, which would require the concomitant alleviation of lymphopenia and neutrophilia. On the other hand, although different stimuli and conditions such as mechanical ventilation, muscle immobilization, severe sepsis, and multiple organ dysfunction as well as neuro/myotoxic agents may contribute to a critical status amongst patients admitted to ICU^47^, all severe patients in our study, with either survivor or deceased outcomes, were under comparable pharmacological and physical management (Tables ST1 and ST2).

A relevant factor associated with late-onset hypogonadism^48^, as well as with an irreversible failure to reinstate testosterone production after critical situations that may compromise the LH-androstenedione axis is old age^49^, which has been linked to senescent dysfunction of Leydig cells^50^. The fact that a majority of non-survivor patients in our study who failed to reinstate testosterone levels are older than 60 years of age would be consistent with the senescence hypothesis. However, our cohort has more patients older than 60 who reinstated their testosterone levels and survived severe COVID-19. Therefore, either Leydig cell senescence only affects a small subset of older patients, or other mechanisms may be invoked to explain failure to restore testosterone production.

Conversely, these observations also suggest that, while the reinstatement of physiological testosterone levels may be mechanistically linked to a return to lymphocyte and neutrophil homeostasis, it may not be required for the relative normalization of other inflammatory pathways, arguably driven by an unmitigated production of IL-6 and other pro-inflammatory cytokines triggered by acute viral infection^51^. In this regard, IL-6 can be produced by a broad range of cells, beyond immune cells^51^, in response, or not, to production of IL-1 β or IL-18 by epithelial or endothelial cells through viral activation of the inflammasome^52^, aided by viral blunting of class I and III interferon pathways^53^. Pro-inflammatory actions of cytokines such as IL-6 are the main inducers of the production of CRP in the liver during infectious and inflammatory processes^54^, and can independently trigger the activation of coagulation pathways, explaining the rise in blood D-dimer levels^51^, as well as complement pathways, which, coupled to direct viral cytopathic effects, explains the elevated circulating levels of tissue damage markers such as LDH and a further exacerbation of a pro-coagulant state^55^. As such, an eventual control of viral replication is expected to lead to a return of these pathways to homeostasis. However, our observations suggest that sufficient and timely resolution of pathogenic hyperinflammation to prevent a lethal outcome may require the additional return to homeostasis of innate and/or adaptive immune cell dynamics and function, possibly assisted in male patients by reinstatement of testosterone production.

There is now a wealth of studies describing the dynamics of immune responses to acute and subacute infection with SARS-CoV-2, including multiparameter and functional analyses of circulating and tissue-associated innate and adaptive immune subpopulations^56,57^. Some of these studies have addressed sex differences in such responses^11,14^. Our analysis in male patients indicates an association of specific immune subpopulations with COVID-19 outcome and a shift of such associations from early (Sample 1) to late (Sample 2) time-points in the course of the disease. For example, the relative representation of terminally differentiated (CD8^+^ TEMRA, CD4^+^ TEMRA) and activated (CD4^+^HLA-DR^+^ and CD8^+^HLA-DR^+^) T cell subpopulations and differentiating B cells modestly correlated with all outcomes except mild disease in Sample 1. This indicates an ongoing early immune response of similar nature and magnitude, regardless of final outcome, as reported by others^56^. In this phase, non-classical monocytes are present in patients with a moderate outcome, while patients with severe survivor and severe deceased outcomes show a predominance of more inflammatory^58^ classical monocytes, in support of a more inflammatory state of these patients, as also evidenced by the clinical biochemical and hematological parameters discussed above.

However, later in the course of disease (Sample 2), a remarkable shift takes place, in particular with regards to correlations with severe survivor as compared to severe deceased patients. As such, while patients with severe survivor outcomes show positive correlations with terminally differentiated (CD4^+^ TEMRA, CD8^+^ TEMRA), activated (HLA-DR^+^ on CD4^+^ and on CD8^+^) and memory (CD4^+^ central memory, CD8^+^ central memory) T cell subpopulations, patients with eventual fatal outcomes evidence a depletion of these subpopulations along with an accumulation of undifferentiated T helper cells (recent thymic emigrant CD4^+^ and naïve CD4^+^). This late shift also affects innate immune populations, as severe survivor patients now correlate with non-classical monocytes over classical monocytes, while the reverse is the case for severe deceased patients. Similar observations have been made by others in studies correlating innate^56^ and adaptive^16^ immune cell subpopulations to COVID-19 outcome.

Importantly, our study additionally correlates relative representations of immune subpopulations to testosterone levels. Thus, higher testosterone levels in Sample 1 are correlated to polarized (Th1, Th17, Th1-17, Th2) and differentiated (CD4^+^ effector memory, CD4^+^ central memory, CD8^+^ TEMRA) T cell subpopulations, and in Sample 2 to a similar range of subpopulations, along with plasmablasts and mature NK cells (CD16^+dim^CD16^+^). These temporal switches in the differentiation profiles of distinct immune subpopulations may suggest that, in patients with lethal outcomes, there may be a defective skewing of T helper cells^38^ and monocytes. A second possible explanation of the apparent depletion of circulating differentiated and polarized cells may be an enhanced clearance or migration from circulation to peripheral tissues^36^. Finally, specific subpopulations may become exhausted in late stages of the disease^39,40^. These three putative mechanisms are not mutually exclusive, and may take place either simultaneously or dynamically at different time points along the clinical course of the patients.

The observed concordance of lethal outcome in male COVID-19 patients with (i) persistent lymphopenia and neutrophilia, (ii) depletion of circulating differentiated T helper and T cytotoxic cells and non-classical monocytes, (iii) accumulation of naïve immune counterparts, and (iv) failure to reinstate physiological levels of testosterone, mirrored by converse phenotypes in severe survivor patients who have undergone equivalent critical illness and management, makes it appealing to hypothesize a mechanistic relationship bonding these coincident phenotypes.

Sex hormones have a profound influence on innate and adaptive immune system development, differentiation and response to challenge^14,17^ More specifically, androgens have a global anti-inflammatory effect^23,59^, reflected in higher frequencies of autoimmune diseases in women or in acquired or genetically determined hypogonadism^18^, as compared to men with a normal XY chromosome complement. On the other hand, testosterone replacement therapy in hypogonadal men attenuates inflammation^60^ and androgens suppress thymic precursor development^61^ and promote the terminal differentiation of T cell subpopulations^62^ and monocyte precursors^63^. Conversely, androgen deprivation through surgical or pharmacological castration in animal models prompts the regeneration of the thymus in aged mice, leading to a relative accumulation of naïve T cell populations (RTE and naïve T cells)^64^ and classical monocytes^65^. A similar effect of androgen deprivation on T cell development and differentiation has been observed in prostate cancer patients, with an expansion of RTE and naive T cells, particularly among CD4^+^ cells^64,66^.

To sum up, the tight association between reinstatement of testosterone and survival from COVID-19 in male patients, along with a reversal of signs of excessive inflammation and immune dysfunction, suggests a functional role for testosterone, beyond being a mere biomarker of outcome, in such recovery.

The limitations of our study include its observational nature on retrospective patients and samples, which has precluded the collection of samples at precisely equivalent time points after symptom onset for all patients. This has most notably impacted longitudinal studies of immune populations which, in the present study, was limited to two temporally separate determinations per male patient studied. A follow-up study is currently ongoing aimed at expanding the present study in this regard, including longitudinal analyses in women. On the other hand, several hypotheses laid out here would require formal testing by means of approaches that are not addressed in the present study. For example, the various mechanisms postulated to explain the depletion of circulating differentiated effector T cells in lethal COVID-19 patients would benefit from additional analyses of senescent and activation states of such populations with appropriate markers, as well as detailed analyses of viral infection and immune cell populations infiltrating key tissues, mainly lung and testis. Finally, pre-clinical animal models would be required for a robust experimental demonstration of mechanistic relationships between testosterone status (e. g., deprivation and replacement) and SARS-CoV-2 infection outcomes, which should also contemplate factors such as age.

## Materials and Methods

### Study design

We studied a group of patients with COVID-19 (249 men and 248 women) admitted at the Vall d’Hebron Hospital between May 1^st^ through June 30^th^ 2020. All patients were diagnosed of SARS-CoV-2 infection by real-time RT-PCR quantitation of viral ORF1a and nucleocapsid RNAs, performed on samples from nasopharyngeal swabs. The frequency distribution relative to age in males and females is illustrated in Extended Data Fig. 1. Co-morbidities considered were: chronic lung disease, cardiovascular disease, diabetes, chronic kidney disease, liver disease, HIV infection with good adherence, obesity (BMI ≥ 30) and cancer. Previously hospitalized patients, recently transplanted, immunosuppressed, and hormonally depleted patients were excluded from the study. A sub cohort of 114 male patients, including patients that entered the intensive care unit (ICU), were selected for longitudinal analysis. The present study was performed with surplus serum samples from routinely tested hospitalized COVID-19 patients, following a protocol reviewed and approved by the Hospital Vall d’Hebron Institutional Review Board (Medical Research Ethics Committee, protocol number PR(AG)329-2020).

Immunophenotyping studies of peripheral blood cells underwent a separate review and approval process (protocol number PR(AG)242/2020).

### Patient classification

Patients were classified as mild, moderate, severe-survivor and severe-deceased as per a 4-point scale (Table ST3).

### Data collection

Clinical data, demographics, co-morbidities, hospital admission, discharge, death dates, time from symptoms onset to hospitalization, length of hospital stay, treatments, requirement for oxygen support, and ICU requirement, were collected from the Hospital database. For reference analyses (497 patients, males and females), data were obtained at (near) hospital admission date. For longitudinal analysis, a sub-cohort (114) of male patients were analyzed throughout the hospitalization.

### Serological determinations

Serum biochemical variables were measured by automated analyzers at the Core Laboratory Facility at the Biochemistry Service of the Vall d’Hebron Hospital. Serum hormones were measured as described in Supplementary Materials. All determinations for the patient population were compared to internal controls used for reference ranges at our core laboratory. Serum hormone levels were determined by chemiluminiscent immunoassays (CLIA) on an AtellicaTM IM Analyzer (Siemens Inc., NY), using Testosterone TSTII (Siemens ref. 10995707) and Luteinizing Hormone (LH) (Siemens ref. 10995634) kits. Androstenedione was measured on a LIASON XL Analyzer (DiaSorin, Saluggia, Italy), using the LiasonR Androstenedione (ref. 318870) assay. As references for healthy men, we used median total serum testosterone levels of 409.72 ng/dL (CI 90 197.44-669.58) for < 50 yrs, and 377.46 ng/dL (CI 90 187.72-684.19) for > 50 years (FDA-approved protocol, https://www.accessdata.fda.gov/cdrh_docs/pdf19/K191533.pdf). For luteinizing hormone (LH), the reference median values were 2.8 mIU/mL (CI 1.5-9.3 mIU/mL) for < 70yrs, and 8.0 mIU/mL (range 3.1-34.6 mIU/mL) for > 70 years. For androstenedione, the reference median value was 1.80 ng/mL (range 0.5-3.5 ng/mL).

### Immuno-phenotyping

Blood was collected in vacutainer tubes containing ethylene-diamine-tetra-acetic acid (EDTA) as anticoagulant (BD-Plymouth, PL6 7BP, UK) and processed within 4 h after collection. Absolute counts and relative numbers of peripheral blood lymphocytes were determined for all study participants using tetra CHROME Tube 1 (CD45-FITC/CD4-PE/CD8-ECD/CD3-PC5) and tetraCHROME Tube 2 (CD45-FITC/CD56-PE/CD19-ECD/CD3-PC5) (Beckman Coulter (BC)) according to the manufacturer’s instructions. Samples were fixed in 1X lysing solution (BC) and acquired on a BC Navios EX instrument.

For multicolor staining and analysis, extended lymphocyte subpopulations were assessed with 5 different flow cytometry panels designed according to the HIPC protocol^67^. Two additional panels were added to analyze the basic lymphocyte populations and RTE. Compensation controls were used in each panel to avoid overlapping of the different fluorochromes. Gating strategies were as described^68^, and the antibody panels are summarized as follows:

Panel 1: General immune phenotype of T, B, and natural killer (NK) lymphocyte subpopulations, gating by CD45 versus SSC.

Panel 2: Gating strategy for differentiated CD4^+^ and CD8^+^ T-cell subsets, based on CD45RA and CCR7 expression defining: CD45RA^+^/CCR7^+^ (naïve), CD45RA^−^/CCR7^+^ (central memory [TCM]), CD45RA^−^/CCR7^−^ (effector memory [TEM]), and CD45RA^+^/CCR7^−^ (terminal effector memory [TEMRA]). CD4^+^ T-helper (Th) populations (Th1, Th2, Th17, Th1–17), based on CCR6 and CXCR3 expression, were analyzed by gating on CD45RA− TCM and TEM cells.

Panel 3: T-regulatory (Treg) cell populations: CD3^+^CD4^+^CD25^+^, CD127^−^, CCR4^+^, and CD45RO^+^.

Panel 4: B-cell populations (naïve, pre-switched, switched memory, and exhausted) depending on expression of IgD and CD27. The differing pattern of CD24^+^ and CD38^+^ expression identified transitional cells and plasmablasts. CD27 and CD21 enabled study of the CD21low population.

Panel 5: Dendritic cells (DC), NK cells, and monocyte populations were analysed in the CD3^−^CD19^−^gate. NK subpopulations (NK dim and NK bright) were studied using CD56 and CD16 expression. The markers CD16 and CD14 were used to identify classical monocytes (CD14^+^CD16^−^) and non-classical monocytes (CD16^+^CD14^−^). DCs were studied selecting the population negative for the following markers: CD3, CD14, CD16, CD19, CD20, and CD56. High expression of HLA-DR and CD11c and CD123 was used to identify plasmacytoid DCs (HLA-DR^+^CD123^+^) and myeloid DCs (HLA-DR^+^CD11c^+^).

Panel 6: Recent thymic emigrant cells (RTEs) were studied using CD3, CD4, CD27, CD31, CD45RA, and CD62L expression.

The composition of mAb mixes, concentrations, clones, and brands are specified in Supplementary Table ST4. The samples were acquired with a NAVIOS EX (BC) flow cytometer. At least 100,000 events were acquired from each sample. Flow cytometry data were analysed with Kaluza Software. Internal quality assurance procedures were performed according to the manufacturer’s instructions. Absolute values were calculated from the absolute number of leucocytes and lymphocytes provided by the haematological analyser (XN-2000; Sysmex, Japan).

### Statistical Analysis

Continuous variables were expressed as mean ± SD or median and interquartile range (IQR). Simple and multiple comparisons were performed using parametric (Student’s t-test or ANOVA) and non-parametric (Mann-Whitney U-test or Kruskal– Wallis) statistical tests with Dunn’s and Tukey’s post hoc tests. Categorical variables were presented as numbers and percentages and compared using the 2-test of the Fisher exact test as appropriate. ROC curves were calculated with the logistic regression model implemented in STATA 14 and the EasyROC web tool (http://www.biosoft.hacettepe.edu.tr/easyROC/), Odd Ratios (OR) were calculated by transforming areas under the curve (AUC) with the Effect Size Converter web tool (https://www.escal.site/). Principal Component Analysis (PCA), multivariate correlation analysis and other calculations, as well as graphic representations, were performed with GraphPad Prism 9.0.2.

## Data Availability

All data described in our manuscript will be made available to interested researchers upon request within appropriate collaboration settings.

## Acknowledgments

The authors wish to thank A. Sánchez Montalvá for advice and the Vall d’Hebron Biochemical Service staff for technical support.

## Author roles

Contributors RP, TMT, ETG and MMG conceived and designed the study, analyzed data, generated figures and tables and wrote the manuscript. ETG, AG, NDT, PGM MLH, FMV and MRB collected clinical samples and data. ETG, MMG, IAM and AG performed experiments, analyzed data and generated figures and tables. MLH, FMV and MRB participated in patient care. RP, TMT, ETG, MMG, PG, MH, RPB, FRF, and RF contributed to data interpretation and critically reviewed the manuscript. All authors approved the final manuscript for submission.

## References

1 Jin, J. M. et al. Gender Differences in Patients With COVID-19: Focus on Severity and Mortality. Front Public Health 8, 152, doi:10.3389/fpubh.2020.00152 (2020).

2 Peckham, H. et al. Male sex identified by global COVID-19 meta-analysis as a risk factor for death and ITU admission. Nat Commun 11, 6317, doi:10.1038/s41467-020-19741-6 (2020).

3 Grasselli, G. et al. Baseline Characteristics and Outcomes of 1591 Patients Infected With SARS-CoV-2 Admitted to ICUs of the Lombardy Region, Italy. JAMA 323, 1574–1581, doi:10.1001/jama.2020.5394 (2020).

4 Karlberg, J., Chong, D. S. & Lai, W. Y. Do men have a higher case fatality rate of severe acute respiratory syndrome than women do? Am J Epidemiol 159, 229–231, doi:10.1093/aje/kwh056 (2004).

5 Noymer, A. & Garenne, M. The 1918 influenza epidemic’s effects on sex differentials in mortality in the United States. Popul Dev Rev 26, 565–581, doi:10.1111/j.1728-4457.2000.00565.x (2000).

6 Price-Haywood, E. G., Burton, J., Fort, D. & Seoane, L. Hospitalization and Mortality among Black Patients and White Patients with Covid-19. N Engl J Med 382, 2534–2543, doi:10.1056/NEJMsa2011686 (2020).

7 Baratchian, M. et al. Androgen regulation of pulmonary AR, TMPRSS2 and ACE2 with implications for sex-discordant COVID-19 outcomes. Scientific Reports 11, 11130, doi:10.1038/s41598-021-90491-1 (2021).

8 Qiao, Y. et al. Targeting transcriptional regulation of SARS-CoV-2 entry factors ACE2 and TMPRSS2. Proc Natl Acad Sci U S A, doi:10.1073/pnas.2021450118 (2020).

9 Caffo, O. et al. Incidence and outcomes of severe acute respiratory syndrome coronavirus 2 infection in patients with metastatic castration-resistant prostate cancer. Eur J Cancer 140, 140–146, doi:10.1016/j.ejca.2020.09.018 (2020).

10 Montopoli, M. et al. Androgen-deprivation therapies for prostate cancer and risk of infection by SARS-CoV-2: a population-based study (N = 4532). Ann Oncol 31, 1040–1045, doi:10.1016/j.annonc.2020.04.479 (2020).

11 Dhindsa, S. et al. Association of Circulating Sex Hormones With Inflammation and Disease Severity in Patients With COVID-19. JAMA Network Open 4, e2111398–e2111398, doi:10.1001/jamanetworkopen.2021.11398 (2021).

12 Rastrelli, G. et al. Low testosterone levels predict clinical adverse outcomes in SARS-CoV-2 pneumonia patients. Andrology 9, 88–98, doi:10.1111/andr.12821 (2021).

13 Salonia, A. et al. Severely low testosterone in males with COVID-19: A case-control study. Andrology, doi:10.1111/andr.12993 (2021).

14 Takahashi, T. et al. Sex differences in immune responses to SARS-CoV-2 that underlie disease outcomes. medRxiv, doi:10.1101/2020.06.06.20123414 (2020).

15 Kreutmair, S. et al. Distinct immunological signatures discriminate severe COVID-19 from non-SARS-CoV-2-driven critical pneumonia. Immunity, doi:10.1016/j.immuni.2021.05.002 (2021).

16 Kuri-Cervantes, L. et al. Comprehensive mapping of immune perturbations associated with severe COVID-19. Sci Immunol 5, doi:10.1126/sciimmunol.abd7114 (2020).

17 Lucas, C. et al. Longitudinal analyses reveal immunological misfiring in severe COVID-19. Nature 584, 463–469, doi:10.1038/s41586-020-2588-y (2020).

18 Klein, S. L. & Flanagan, K. L. Sex differences in immune responses. Nat Rev Immunol 16, 626–638, doi:10.1038/nri.2016.90 (2016).

19 Cephus, J. Y. et al. Testosterone Attenuates Group 2 Innate Lymphoid Cell-Mediated Airway Inflammation. Cell Rep 21, 2487–2499, doi:10.1016/j.celrep.2017.10.110 (2017).

20 Laffont, S. & Guery, J. C. Deconstructing the sex bias in allergy and autoimmunity: From sex hormones and beyond. Adv Immunol 142, 35–64, doi:10.1016/bs.ai.2019.04.001 (2019).

21 Sette, A. & Crotty, S. Adaptive immunity to SARS-CoV-2 and COVID-19. Cell 184, 861–880, doi:10.1016/j.cell.2021.01.007 (2021).

22 Wang, C. et al. Sex differences in group 2 innate lymphoid cell-dominant allergic airway inflammation. Mol Immunol 128, 89–97, doi:10.1016/j.molimm.2020.09.019 (2020).

23 Furman, D. et al. Systems analysis of sex differences reveals an immunosuppressive role for testosterone in the response to influenza vaccination. Proc Natl Acad Sci U S A 111, 869–874, doi:10.1073/pnas.1321060111 (2014).

24 Vom Steeg, L. G. et al. Age and testosterone mediate influenza pathogenesis in male mice. Am J Physiol Lung Cell Mol Physiol 311, L1234–L1244, doi:10.1152/ajplung.00352.2016 (2016).

25 Zhang, J., Yu, M., Tong, S., Liu, L. Y. & Tang, L. V. Predictive factors for disease progression in hospitalized patients with coronavirus disease 2019 in Wuhan, China. J Clin Virol 127, 104392, doi:10.1016/j.jcv.2020.104392 (2020).

26 Huang, C. et al. Clinical features of patients infected with 2019 novel coronavirus in Wuhan, China. Lancet 395, 497–506, doi:10.1016/S0140-6736(20)30183-5 (2020).

27 Laguna-Goya, R. et al. IL-6-based mortality risk model for hospitalized patients with COVID-19. J Allergy Clin Immunol 146, 799–807 e799, doi:10.1016/j.jaci.2020.07.009 (2020).

28 Hu, J., Zhang, Z., Shen, W. J. & Azhar, S. Cellular cholesterol delivery, intracellular processing and utilization for biosynthesis of steroid hormones. Nutr Metab (Lond) 7, 47, doi:10.1186/1743-7075-7-47 (2010).

29 Teblick, A., Langouche, L. & Van den Berghe, G. Anterior pituitary function in critical illness. Endocr Connect 8, R131–R143, doi:10.1530/EC-19-0318 (2019).

30 Malkin, C. J. et al. The effect of testosterone replacement on endogenous inflammatory cytokines and lipid profiles in hypogonadal men. J Clin Endocrinol Metab 89, 3313–3318, doi:10.1210/jc.2003-031069 (2004).

31 Guo, J. et al. The adult human testis transcriptional cell atlas. Cell Res 28, 1141–1157, doi:10.1038/s41422-018-0099-2 (2018).

32 Hikmet, F. et al. The protein expression profile of ACE2 in human tissues. Mol Syst Biol 16, e9610, doi:10.15252/msb.20209610 (2020).

33 Yang, M. et al. Pathological Findings in the Testes of COVID-19 Patients: Clinical Implications. Eur Urol Focus 6, 1124–1129, doi:10.1016/j.euf.2020.05.009 (2020).

34 Kvedaraite, E. et al. Major alterations in the mononuclear phagocyte landscape associated with COVID-19 severity. Proceedings of the National Academy of Sciences 118, e2018587118, doi:10.1073/pnas.2018587118 (2021).

35 Gatti, A., Radrizzani, D., Viganò, P., Mazzone, A. & Brando, B. Decrease of Non-Classical and Intermediate Monocyte Subsets in Severe Acute SARS-CoV-2 Infection. Cytometry Part A 97, 887-890, doi:https://doi.org/10.1002/cyto.a.24188 (2020).

36 Chen, Z. & John Wherry, E. T cell responses in patients with COVID-19. Nature Reviews Immunology 20, 529–536, doi:10.1038/s41577-020-0402-6 (2020).

37 Gil-Etayo, F. J. et al. T-Helper Cell Subset Response Is a Determining Factor in COVID-19 Progression. Frontiers in Cellular and Infection Microbiology 11, doi:10.3389/fcimb.2021.624483 (2021).

38 Brooks, D. G., Teyton, L., Oldstone, M. B. A. & McGavern, D. B. Intrinsic Functional Dysregulation of CD4 T Cells Occurs Rapidly following Persistent Viral Infection. Journal of Virology 79, 10514-10527, doi:doi:10.1128/JVI.79.16.10514-10527.2005 (2005).

39 Diao, B. et al. Reduction and Functional Exhaustion of T Cells in Patients With Coronavirus Disease 2019 (COVID-19). Frontiers in Immunology 11, doi:10.3389/fimmu.2020.00827 (2020).

40 Kusnadi, A. et al. Severely ill COVID-19 patients display impaired exhaustion features in SARS-CoV-2-reactive CD8(+) T cells. Sci Immunol 6, doi:10.1126/sciimmunol.abe4782 (2021).

41 Knight, S. R. et al. Risk stratification of patients admitted to hospital with covid-19 using the ISARIC WHO Clinical Characterisation Protocol: development and validation of the 4C Mortality Score. BMJ 370, m3339, doi:10.1136/bmj.m3339 (2020).

42 Bastard, P. et al. Autoantibodies against type I IFNs in patients with life-threatening COVID-19. Science 370, eabd4585, doi:10.1126/science.abd4585 (2020).

43 Chakravarty, D. et al. Sex differences in SARS-CoV-2 infection rates and the potential link to prostate cancer. Commun Biol 3, 374, doi:10.1038/s42003-020-1088-9 (2020).

44 Meng, Y. et al. Sex-specific clinical characteristics and prognosis of coronavirus disease-19 infection in Wuhan, China: A retrospective study of 168 severe patients. PLoS Pathog 16, e1008520, doi:10.1371/journal.ppat.1008520 (2020).

45 Ten-Caten, F. et al. In-depth analysis of laboratory parameters reveals the interplay between sex, age, and systemic inflammation in individuals with COVID-19. Int J Infect Dis 105, 579–587, doi:10.1016/j.ijid.2021.03.016 (2021).

46 Venet, F. et al. Longitudinal assessment of IFN-I activity and immune profile in critically ill COVID-19 patients with acute respiratory distress syndrome. Critical Care 25, 140, doi:10.1186/s13054-021-03558-w (2021).

47 Friedrich, O. et al. The Sick and the Weak: Neuropathies/Myopathies in the Critically Ill. Physiol Rev 95, 1025–1109, doi:10.1152/physrev.00028.2014 (2015).

48 Wu, F. C. et al. Identification of late-onset hypogonadism in middle-aged and elderly men. N Engl J Med 363, 123–135, doi:10.1056/NEJMoa0911101 (2010).

49 Mulligan, T., Iranmanesh, A. & Veldhuis, J. D. Pulsatile iv infusion of recombinant human LH in leuprolide-suppressed men unmasks impoverished Leydig-cell secretory responsiveness to midphysiological LH drive in the aging male. J Clin Endocrinol Metab 86, 5547–5553, doi:10.1210/jcem.86.11.8004 (2001).

50 Zhang, C. et al. FOXO4-DRI alleviates age-related testosterone secretion insufficiency by targeting senescent Leydig cells in aged mice. Aging (Albany NY) 12, 1272–1284, doi:10.18632/aging.102682 (2020).

51 Velazquez-Salinas, L., Verdugo-Rodriguez, A., Rodriguez, L. L. & Borca, M. V. The Role of Interleukin 6 During Viral Infections. Front Microbiol 10, 1057, doi:10.3389/fmicb.2019.01057 (2019).

52 Yap, J. K. Y., Moriyama, M. & Iwasaki, A. Inflammasomes and Pyroptosis as Therapeutic Targets for COVID-19. J Immunol 205, 307–312, doi:10.4049/jimmunol.2000513 (2020).

53 Schultze, J. L. & Aschenbrenner, A. C. COVID-19 and the human innate immune system. Cell 184, 1671–1692, doi:10.1016/j.cell.2021.02.029 (2021).

54 Tanaka, T., Narazaki, M. & Kishimoto, T. IL-6 in inflammation, immunity, and disease. Cold Spring Harb Perspect Biol 6, a016295, doi:10.1101/cshperspect.a016295 (2014).

55 de Nooijer, A. H. et al. Complement Activation in the Disease Course of Coronavirus Disease 2019 and Its Effects on Clinical Outcomes. J Infect Dis 223, 214–224, doi:10.1093/infdis/jiaa646 (2021).

56 Carsetti, R. et al. Different Innate and Adaptive Immune Responses to SARS-CoV-2 Infection of Asymptomatic, Mild, and Severe Cases. Front Immunol 11, 610300, doi:10.3389/fimmu.2020.610300 (2020).

57 Zhang, J. Y. et al. Single-cell landscape of immunological responses in patients with COVID-19. Nat Immunol 21, 1107–1118, doi:10.1038/s41590-020-0762-x (2020).

58 Kapellos, T. S. et al. Human Monocyte Subsets and Phenotypes in Major Chronic Inflammatory Diseases. Frontiers in Immunology 10, doi:10.3389/fimmu.2019.02035 (2019).

59 Gubbels Bupp, M. R. & Jorgensen, T. N. Androgen-Induced Immunosuppression. Front Immunol 9, 794, doi:10.3389/fimmu.2018.00794 (2018).

60 Corrales, J. J. et al. Enhanced immunological response by dendritic cells in male hypogonadism. Eur J Clin Invest 42, 1205–1212, doi:10.1111/j.1365-2362.2012.02712.x (2012).

61 Lai, J. J. et al. Androgen receptor influences on body defense system via modulation of innate and adaptive immune systems: lessons from conditional AR knockout mice. Am J Pathol 181, 1504–1512, doi:10.1016/j.ajpath.2012.07.008 (2012).

62 Brown, M. A. & Su, M. A. An Inconvenient Variable: Sex Hormones and Their Impact on T Cell Responses. J Immunol 202, 1927–1933, doi:10.4049/jimmunol.1801403 (2019).

63 Consiglio, C. R. & Gollnick, S. O. Androgen Receptor Signaling Positively Regulates Monocytic Development. Frontiers in Immunology 11, doi:10.3389/fimmu.2020.519383 (2020).

64 Sutherland, J. S. et al. Activation of Thymic Regeneration in Mice and Humans following Androgen Blockade. The Journal of Immunology 175, 2741–2753, doi:10.4049/jimmunol.175.4.2741 (2005).

65 van Dommelen, S. L. et al. Regeneration of dendritic cells in aged mice. Cell Mol Immunol 7, 108–115, doi:10.1038/cmi.2009.114 (2010).

66 Morse, M. D. & McNeel, D. G. Prostate cancer patients on androgen deprivation therapy develop persistent changes in adaptive immune responses. Hum Immunol 71, 496–504, doi:10.1016/j.humimm.2010.02.007 (2010).

67 Maecker, H. T., McCoy, J. P. & Nussenblatt, R. Standardizing immunophenotyping for the Human Immunology Project. Nat Rev Immunol 12, 191–200, doi:10.1038/nri3158 (2012).

68 Garcia-Prat, M. et al. Extended immunophenotyping reference values in a healthy pediatric population. Cytometry B Clin Cytom 96, 223–233, doi:10.1002/cyto.b.21728 (2019).

