## Supplemental Files for "Recovery of serum testosterone levels is an accurate predictor of survival from COVID-19 in male patients"

**Table ST1. Treatment comparison by outcome in male patients.**

| Treatments | Total 249 (%) | Mild-Moderate 114 (%) | Severe 135 (%) | p-value |
|---|---|---|---|---|
| Hydroxychloroquine | 241 (96.79) | 112 (98.24) | 129 (95.55) | 0.2953 |
| Antibiotics | 238 (95.58) | 104 (91.23) | 134 (99.26) | 0.0031 |
| Antivirals | 196 (78.71) | 93 (81.58) | 103 (76.30) | 0.3528 |
| Corticoids | 66 (26.5) | 13 (11.40) | 53 (39.26) | <0.0001 |
| Immunomodulators | 130 (52.21) | 23 (20.17) | 107 (79.26) | <0.0001 |
| Anticoagulants | 123 (49.40) | 34 (29.82) | 89 (65.92) | <0.0001 |
| Analgesics | 28 (11.24) | 13 (11.40) | 15 (11.11) | 0.9999 |

**Table ST2. Treatment comparison by outcome in female patients.**

| Treatments | Total 248 (%) | Mild-Moderate 145 (%) | Severe 103 (%) | P-value |
|---|---|---|---|---|
| Hydroxychloroquine | 228 (91.94) | 133 (91.72) | 95 (92.23) | 0.9999 |
| Antibiotics | 231 (93.15) | 128 (88.28) | 103 (100.00) | 0.0001 |
| Antivirals | 215 (86.69) | 129 (88.97) | 86 (83.5) | 0.2557 |
| Corticosteroids | 41 (16.53) | 11 (7.59) | 30 (29.13) | <0.0001 |
| Immunomodulators | 95 (38.31) | 23 (15.86) | 72 (69.90) | <0.0001 |
| Anticoagulants | 70 (28.23) | 10 (6.90) | 60 (58.25) | <0.0001 |
| Analgesics | 65 (26.21) | 40 (27.59) | 25 (24.27) | 0.6605 |

**Table ST3. WHO classification of disease outcome** (adapted from Grein *et al.*[1])

| Group | Outcome | Definition | Stay Type |
|---|---|---|---|
| **1** | Mild | Not hospitalized or hospitalized without oxygen | Discharge from emergency to home or Ward |
| **2** | Moderate | Hospitalized with low flow oxygen by mask or nasal prongs or with high flow oxygen | Ward |
| **3** | Severe-Survivor | Hospitalized with non-invasive ventilation or with invasive mechanical ventilation | ICU |
| **4** | Severe-Deceased | Death | Exitus |

1. Grein J, Ohmagari N, Shin D*, et al.* Compassionate Use of Remdesivir for Patients with Severe Covid-19. *N Engl J Med* 2020; **382**(24): 2327-36. 10.1056/NEJMoa2007016.

### Table ST4. Panels and antibodies used for immunophenotyping.

| General lymphocyte populations | Fluorophore | Isotype | Clone |
|---|---|---|---|
| CD45/CD8/CD4/CD3 | FITC/PE/ECD/PC5 | IgG2b/IgG1/IgG1/IgG1 | B3821F4A/SFCI12T4D11/SFCI21Thy2D3/UCHT1 |
| CD45/CD56/CD19/CD3 | FITC/PE/EDC/PC5 | IgG2b/IgG1/IgG1/IgG1 | B3821F4A/SFCI12T4D11/SFCI21Thy2D3/UCHT1 |

| T-cell populations | | | |
|---|---|---|---|
| CXCR3/CD183 | AF488 | IgG1 | G025H7 |
| CCR7/CD197 | PE | IgG2a | G043H7 |
| CD45RA | ECD | IgG1 | ALB11 |
| CCR6/CD196 | PC7 | IgG2a | B-R35 |
| CD4 | APC | IgG1 | 13B8.2 |
| CD8 | APC700 | IgG1 | SFCI21Thy2D3 (T8) |
| CD3 | APC750 | IgG1 | UCHT1 |
| HLA-DR | PB | IgG1 | Immu-357 |
| CD45 | KRO | IgG1 | J.33 |

| Recent Thymic Emigrant | | | |
|---|---|---|---|
| CD31 | FITC | IgG1 | 5.6E |
| CD62L | PE | IgG1 | DREG56 |
| CD3 | ECD | IgG1 | UCHT1 |
| CD27 | PC7 | IgG1 | 1A4CD27 |
| CD4 | APC | IgG1 | 13B8.2 |
| CD45RA | PB | IgG1 | 2H4LDH11LDB9 (2H4) |
| CD45 | KRO | IgG1 | J.33 |

| T regulatory cell population | | | |
|---|---|---|---|
| CD45RO | FITC | IgG2a | UCHL1 |
| CD25 | PE | IgG2a | B1.49.9 |
| CD3 | ECD | IgG1 | UCHT1 |
| CCR4/ CD194 | PC7 | IgG1 | 1G1 |
| CD4 | APC | IgG1 | 13B8.2 |
| CD127 | APC700 | IgG1 | R34.34 |
| HLA-DR | PB | IgG1 | Immu-357 |
| CD45 | KRO | IgG1 | J.33 |

| DC/Monocytes/NK | | | |
|---|---|---|---|
| CD16 | FITC | IgG1 | 3G8 |
| CD11c | PE | IgG1 | BU15 |
| CD3 | ECD | IgG1 | UCHT1 |
| CD19 | ECD | IgG1 | J3-119 |
| CD20 | ECD | IgG2a | B9E9(HRC20) |
| CD56 | PC7 | IgG1 | N901 (NKH-1) |
| CD123 | APC | IgG1 | SSDCLY107D2 |
| CD14 | APC750 | IgG1 | RMO52 |
| HLA-DR | PB | IgG1 | Immu-357 |

| B-cell populations | | | |
|---|---|---|---|
| IgD | FITC | IgG2a | IA6-2 |
| CD21 | PE | IgG1 | BL13 |
| CD19 | ECD | IgG1 | J3.119 |
| CD27 | PC7 | IgG1 | 1A4CD27 |
| CD24 | APC | IgG1 | ALB9 |
| CD38 | APC750 | IgG1 | LS198-4-3 |
| IgM | PB | IgG1 | SA-DA4 |
| CD45 | KRO | IgG1 | J.33 |

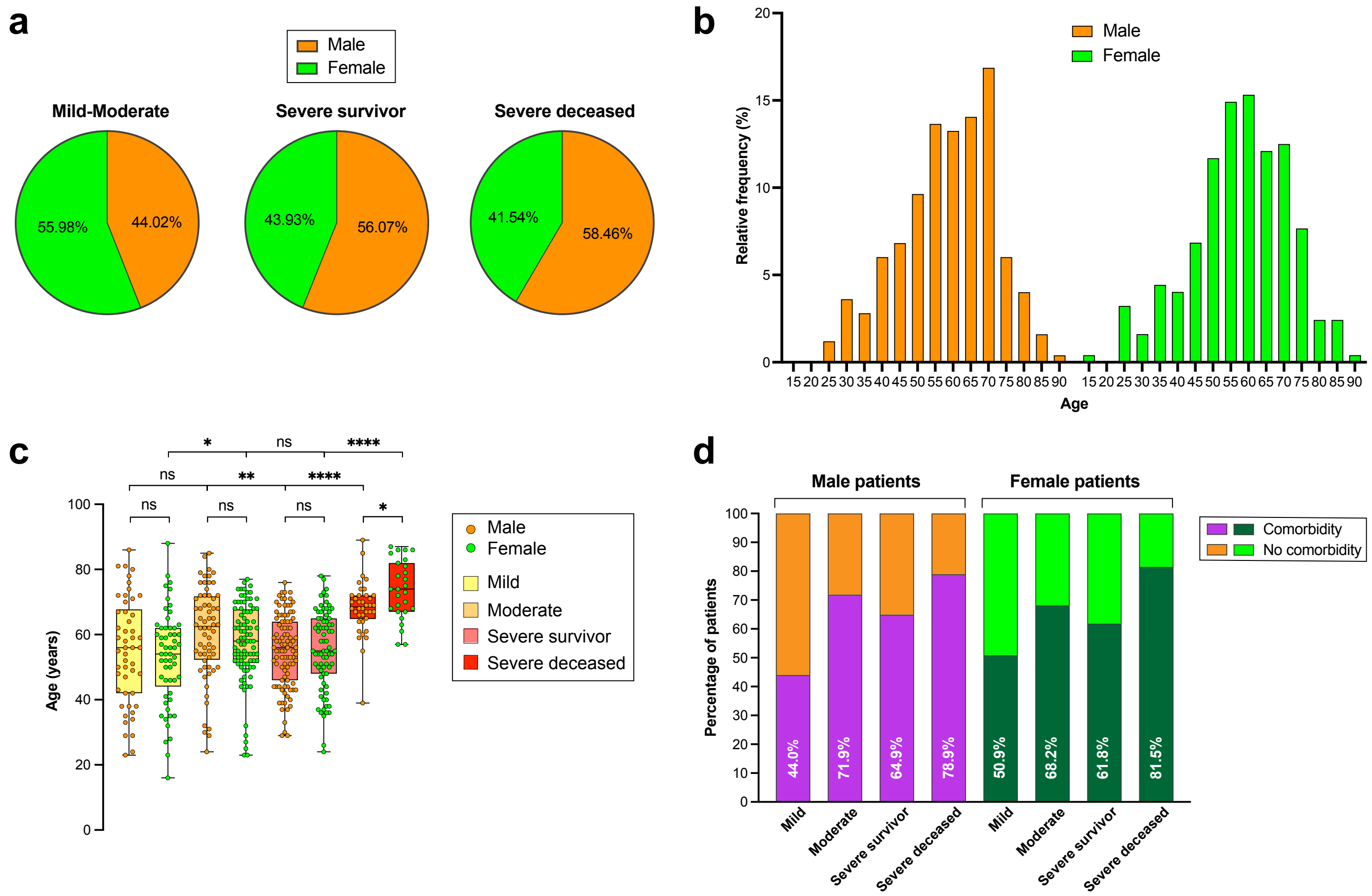


**Figure SF1 (linked to Tables 1 and 2)**. Patient distributions by outcome (**a**), age (**b**), age and outcome (**c**) and comorbidities and outcome (**d**).

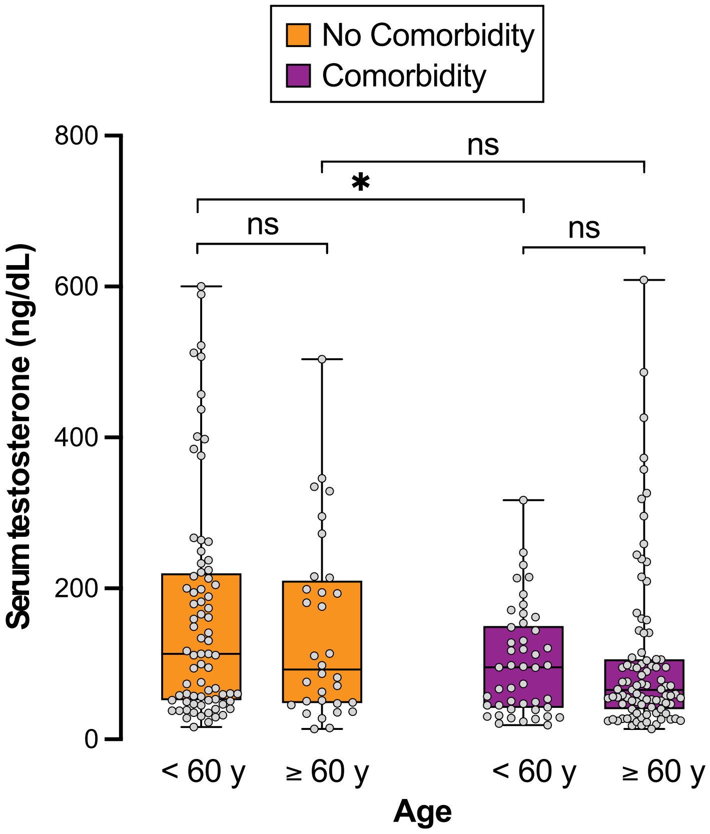


**Figure SF2 (linked to Figure 2)**. Distribution of male patients with comorbidities according to age.

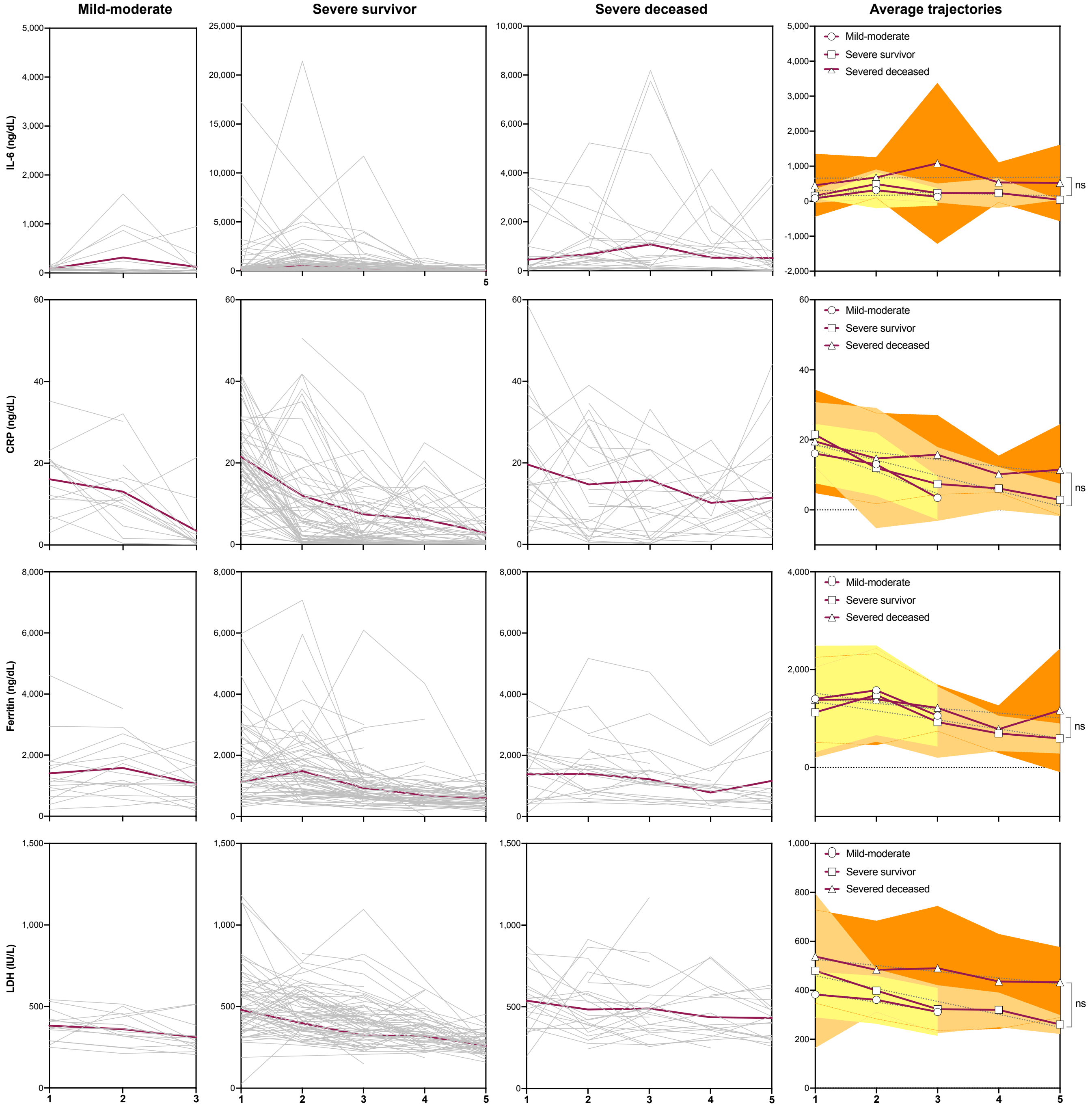


**Figure SF3 (linked to Figure 3)**. Longitudinal analysis in male patients of serum levels of IL-6, C-reactive protein (CRP), ferritin and lactate dehydrogenase (LDH). For severe survivor and severe deceased outcomes, the trajectories of longitudinal determinations were submitted to linear regression analysis, and the resulting slopes compared for significance by two-way ANOVA. ns denotes not significant.

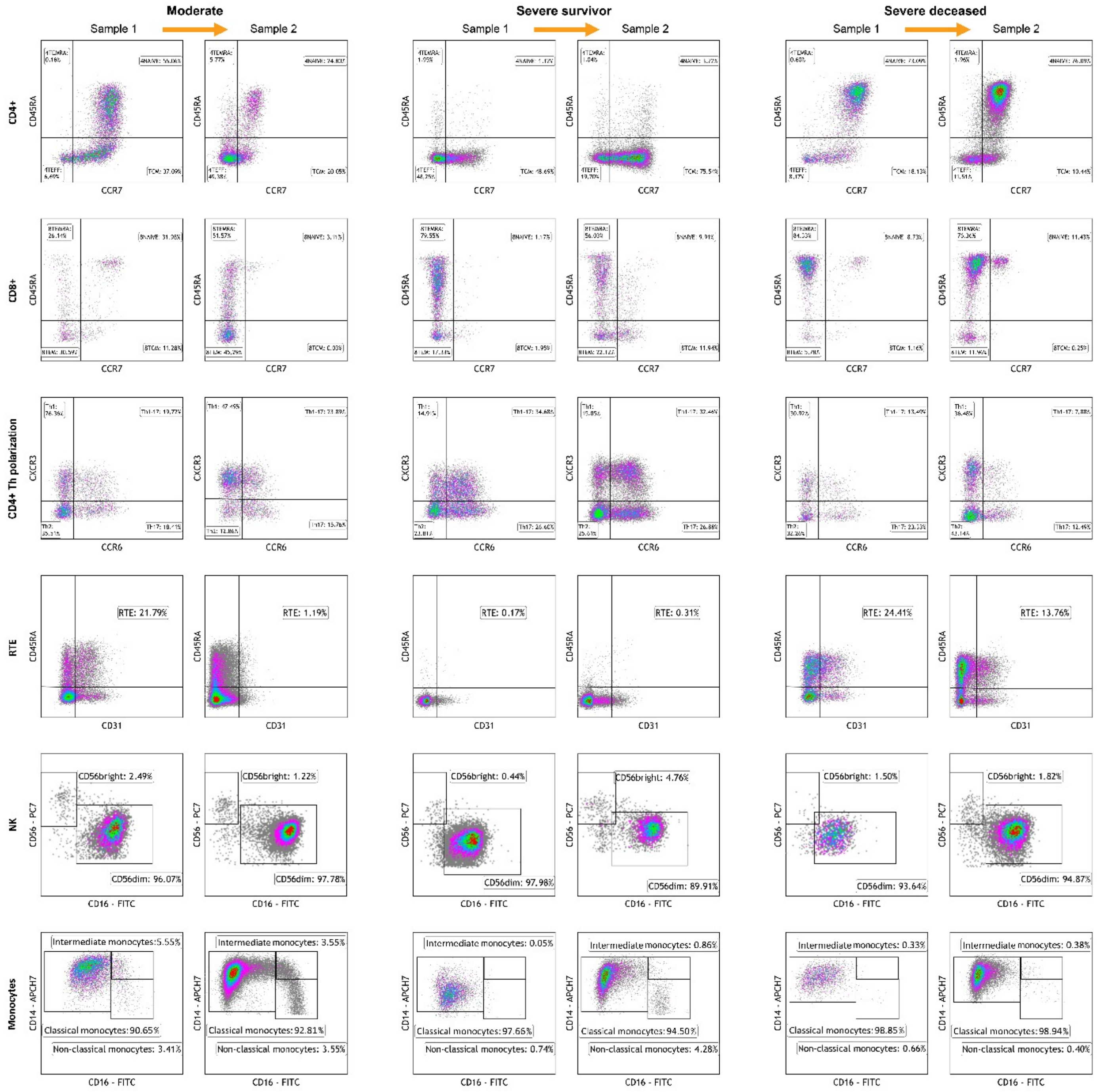


**Figure SF4 (linked to Figure 5)**. Flow cytometry analysis of circulating immune subpopulations in three illustrative cases with moderate, severe survivor and severe deceased outcomes. For each case, Sample 1 corresponds to the first available analysis post-admission, and Sample 2 corresponds to a subsequent analysis, separated from Sample 1 by 6-12 days. CD4+ and CD8+ subpopulations were further segmented into the indicated subpopulations by analyzing for CD45RA and CCR7 expression. CD3+ subpopulations were further segmented into subpopulations by analyzing the expression of the polarization markers CXCR3 and CCR6. Recent thymic emigrant (RTE) populations were identified and scored through the expression of CD3, CD45RA and CD31. Natural killer (NK) cell subpopulations were segmented by analyzing the expression of CD56 and CD16. Monocyte subpopulations were segmented by analyzing the expression of CD14 and CD16. Not shown are B cell and dendritic cell subpopulations, which did not show significant shifts in repertoire between outcomes and longitudinal sampling. 4TEFF, 4TCM, 4TEMRA, 8TEFF, 8TCM, 8TEMRA: CD4+ or CD8+ effector, central memory or T effector memory re-expressing CD45RA, respectively.
